# Zorro versus Covid-19: fighting the pandemic with face masks

**DOI:** 10.1101/2021.01.04.20237578

**Authors:** Olivier Damette

## Abstract

To confront the global Covid-19 pandemic and reduce the spread of the virus, we need to better understand if face mask use is effective to contain the outbreak and investigate the potential drivers in favor of mask adoption. It is highly questionable since there is no consensus among the general public despite official recommendations. For the first time, we conduct a panel econometric exercise to assess the dynamic impact of face mask use on both infected cases and fatalities at a global scale. We reveal a negative impact of mask wearing on fatality rates and on the Covid-19 number of infected cases. The delay of action varies from around 7 days to 28 days concerning infected cases but is more longer concerning fatalities. We also document the increasing adoption of mask use over time. We find that population density and pollution levels are significant determinants of heterogeneity regarding mask adoption across countries, while altruism, trust in government and demographics are not. Surprisingly, government effectiveness and income level (GDP) have an unexpected influence. However, strict government policies against Covid-19 have the most significant effect on mask use. Therefore, the most effective way of increasing the level of mask wearing is to enforce strict laws on the wearing of masks.

---

To confront the global Covid-19 pandemic and reduce the spread of the virus, we need to improve our understanding of the factors that influence its spread. Many governments and public health departments have promoted and imposed various mitigation measures to contain the spread of Covid-19 (1–2); indeed, such measures are crucial in current times when there is no vaccine available. The heterogeneity in fatality rates across the globe reflects differences in how well countries have managed the pandemic (3), the effectiveness of the various policies, and the extent to which they have been promoted by public authorities and adopted by populations.

One of the most widely debated of such policies, especially among the general public, is the wearing of face masks. While face mask use is encouraged by most governments, support for their use has been limited among general populations (4). The effectiveness of masks in reducing the transmission of Covid-19 has been contested, and levels of self-reported mask use differ considerably between countries (6-7).

This absence of consensus among general populations can be explained not only by differences in individuals’ subjective concerns (7) but also by conflicting national guidelines and public communication. The latter is a crucial area of concern since political leaders’ words and actions affect people’s behavior (8); people need official guidance (9). However, public guidance has been subject to much fluctuation and is characterized by a lack of consistency among political leaders. President Trump in the US is a prime example of a leader whose opinions on this issue have been inconsistent. Likewise, the French government did not support face mask use last March, considering it to be ineffective, but changed its guidance to support the wearing of face masks in late May. Even the World Health Organization initially advised against widespread mask wearing before changing its recommendation on June, 5, 2020 (10-11). Beyond individual-level variations, heterogeneity between countries still exists; mask wearing has been recommended since the early stages of the pandemic by governments in East and South East Asia. It was only later recommended in many Western countries and is still not recommended at all in some Nordic countries. Policies on face mask use are also more strictly enforced in Asian countries. The rationale choice for this is that a voluntary policy would likely lead to insufficient compliance and would, hence, be less effective than a mandatory policy (12).

However, the effectiveness of mask wearing policies to reduce the spread of the Covid-19 virus, especially when they go beyond the precautionary principle, has been the subject of much scientific debate (7). Studies on the role face masks can play in mitigating the pandemic are scarce, particularly statistical and global-scale studies. This is partly due to limited data at the level of countries and the difficulty of conducting clinical trials (which may be regarded as exploiting vulnerable populations) at the individual level.

A very small body of literature has found that mandatory and voluntary mask policies should mitigate the transmission and, thus, the spread of the Covid-19 pandemic. The effects of face masks have been examined in a large survey (14) and a meta-analysis (15). Both studies concluded that face mask use reduces the transmission of infected droplets in both laboratory and clinical contexts. In addition, public mask use is most effective in reducing the transmission of the virus when compliance is high and when there is a high level of trust in politicians (5). While clinical masks are the best protective solution, surgical and comparable cloth masks have also been shown to reduce transmission, albeit in a less comprehensive way (14).

Experimental studies on animals confirm the effectiveness of mask use: protected animals were found to be less infected and sick than their mask-free neighbors (15). Results based on mathematical models (16–17) show that face mask use by the public could make a major contribution to reducing the impact of the Covid-19 pandemic. If masks were used in public all the time (not just from when symptoms first appear), the effective reproduction number could be reduced to less than one (16). From a theoretical and mathematical perspective, face masks, even if they have only a limited protective effect, can reduce the total number of infections and deaths and delay the peak of the epidemic (4).

A small number of recent statistical studies have confirmed these results. Based on linear correlation and projections exercises and using data from the US, Italy, and Wuhan city in China, one study concluded that wearing a face mask in public is the most effective means of preventing Covid-19 transmission (17). However, potential biases, especially regarding the failure to control for other non-pharmaceutical policies, have been reported. Very recently, an econometric study demonstrated the positive results of introducing masks for employees in US states in April 2020 (19). Likewise, in Germany, the introduction of mandatory mask wearing has reduced the growth rate of Covid-19 cases. The study exploits the fact that the obligation to wear face masks in public transport, shops, and workplaces was introduced much earlier in Jena area (on 6th April) than in all other regions in Germany (around 20 days later). Generally, these findings are in agreement with the assumptions of epidemiologists and virologists regarding the benefits of reducing virus particle transmission by wearing a face mask (20–21).

Little information is available on a global scale. However, an analysis of the socio-economic determinants of Covid-19 mortality across countries demonstrated that a longer duration of mask wearing by the public was negatively associated with mortality (10). In this study, the effect of mask wearing is proxied by the delay between the first infected case and a government recommendation on mask wearing by the public. This study finds that ‘in countries with public policies and cultural norms supporting public mask-wearing, per-capita coronavirus mortality increased on average by just 15.8% each week, as compared with 62.1% each week in other countries’.

Concerning the global cross-country determinants that influence attitudes to face masks, a small number of studies have been conducted on the level of individuals in Germany based on survey data (22) and Facebook Health Behavior Survey conducted in eight industrialized countries (7). The first one found that worries about the current pandemic have the largest positive influence on mask wearing. Self-protection, protection of others, and perceptions of others’ judgment – especially for young people – have also been found to be significant drivers. Demographic factors (e.g., age, gender) were not found to be significant drivers. The aforementioned study found that self-reported mask use differed considerably across the eight countries considered. In contrast to previous findings, sex- and age-specific patterns about threat perception, confidence in the healthcare system, and the likelihood of adopting preventive behaviors were also documented (6). Older individual-level studies on the severe acute respiratory syndrome (SARS) epidemic of 2003 in Hong Kong (23) reported that women, people in the 50–59 age group, and married respondents were more likely to wear face masks. Some other studies of different but related nature analysed the practices about face masks/N95 respirators utilization in Poland (24) whereas (25) investigated the social and behavioral consequences of mask policies face with the COVID-19 pandemic using 7000 German participants from April, 14 to May, 26, 2020. They reveal that mandatory policy is more effective than voluntary policy.

To the best of our knowledge, we present here the first statistical analysis of the effectiveness of mask wearing to reduce the spread of Covid-19 at a global scale. 1) We compute dynamic panel econometric estimates to assess the impact of mask wearing on both infections and fatality rates per capita. Although some data is available on the number of tests being conducted, no official data on mask wearing exists. Previous studies used a dummy variable by taking into account the date of mandatory mask policy introduction and the duration variable (10). For this study, we collected individual data from the Covid-19 World Survey Data API jointly conducted by the University of Maryland and Facebook (26). We obtained estimates of the percentage of people in a given country that used face masks daily from April, 23 to July, 15, 2020. We developed a dynamic model that included lagged effects to control for the dynamics of the epidemic over time, including delays between infection and case confirmation and incubation period. Our dynamic model indirectly accounts for potential simultaneous impacts of other determinants of Covid-19 outbreak. We also included additional controls to directly account for certain factors that may have influenced viral transmission (e.g., mobility, temperatures). Finally, we controlled for the effects of other non-pharmaceutical mitigation measures during the studied period by adding the number of Covid-19 tests (testing policy) and a stringency index (reflecting all mitigation measures) as additional explanatory variables. 2) We then performed a cross-sectional statistical analysis to examine the socio-economic determinants of mask wearing across countries. For this analysis, we examined a different set of determinants to understand the heterogeneities regarding mask adoption across countries and the factors that can increase the number of individuals that wear a face mask.

## Is mask wearing really effective on a global scale?

We first conducted an econometric exercise for 96 countries for the period April, 23 to July, 15, 2020. To account for lags in epidemic dynamics, we collected data on infected cases and fatalities for a longer period from January, 1, 2020. Our global panel data covers all parts of the globe, but we narrow this down to European countries for robustness checks.

Our dynamic panel data estimates (Mean Group here) reveal that masks wearing has a clear (P ¡ 0.01) negative impact on infected cases with a 7- to 28-day lag but that this negative effect disappears after 42 days (Table 1). The most significant negative effect is obtained after a lag of around 27 days (Fig. 1). After 32 days (the gray area in Fig. 1), the coefficient associated with the mask use variable becomes insignificant.

**Table 1:**
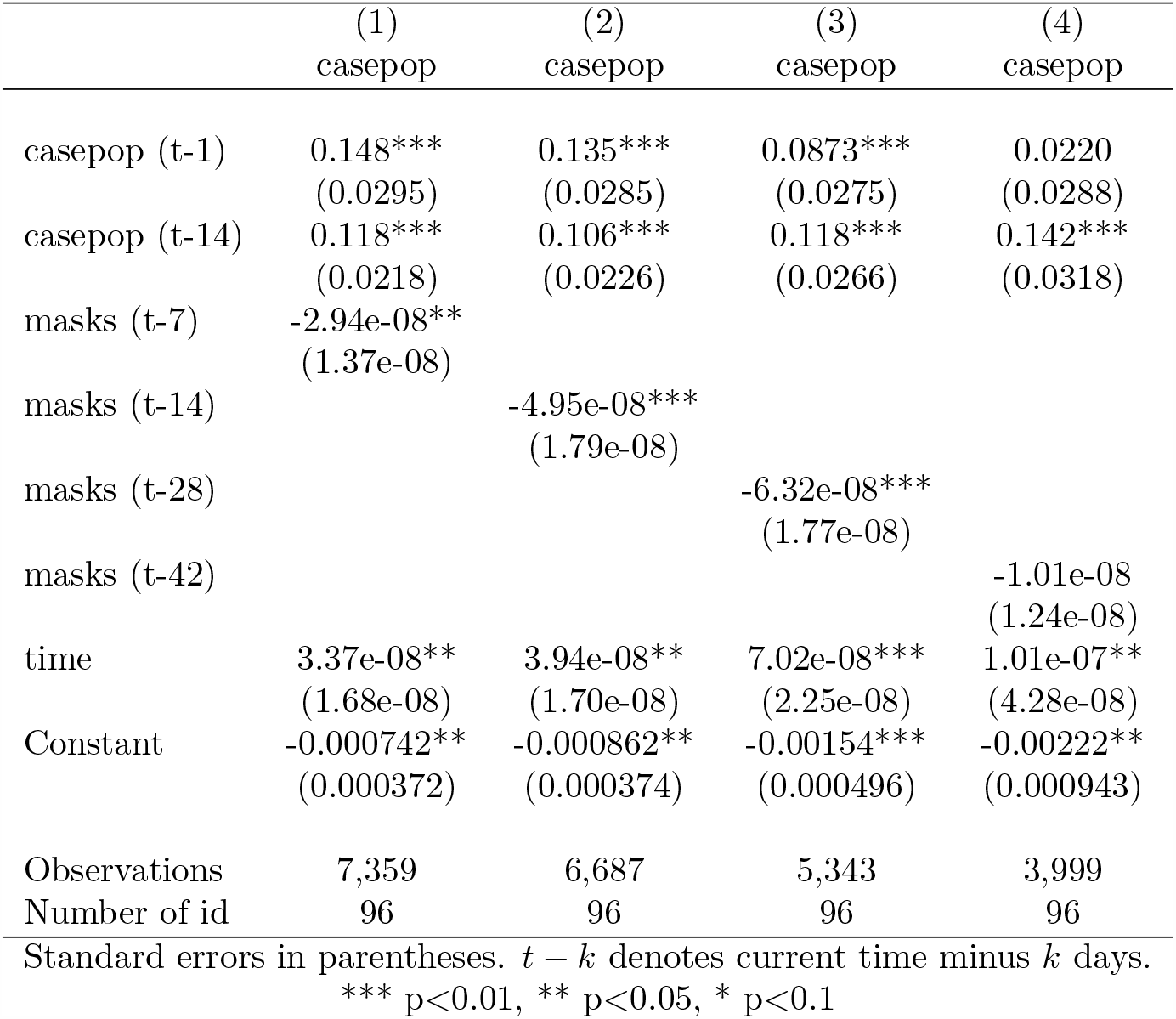
Masks effects on Covid-19 infected cases.

**Figure 1:**
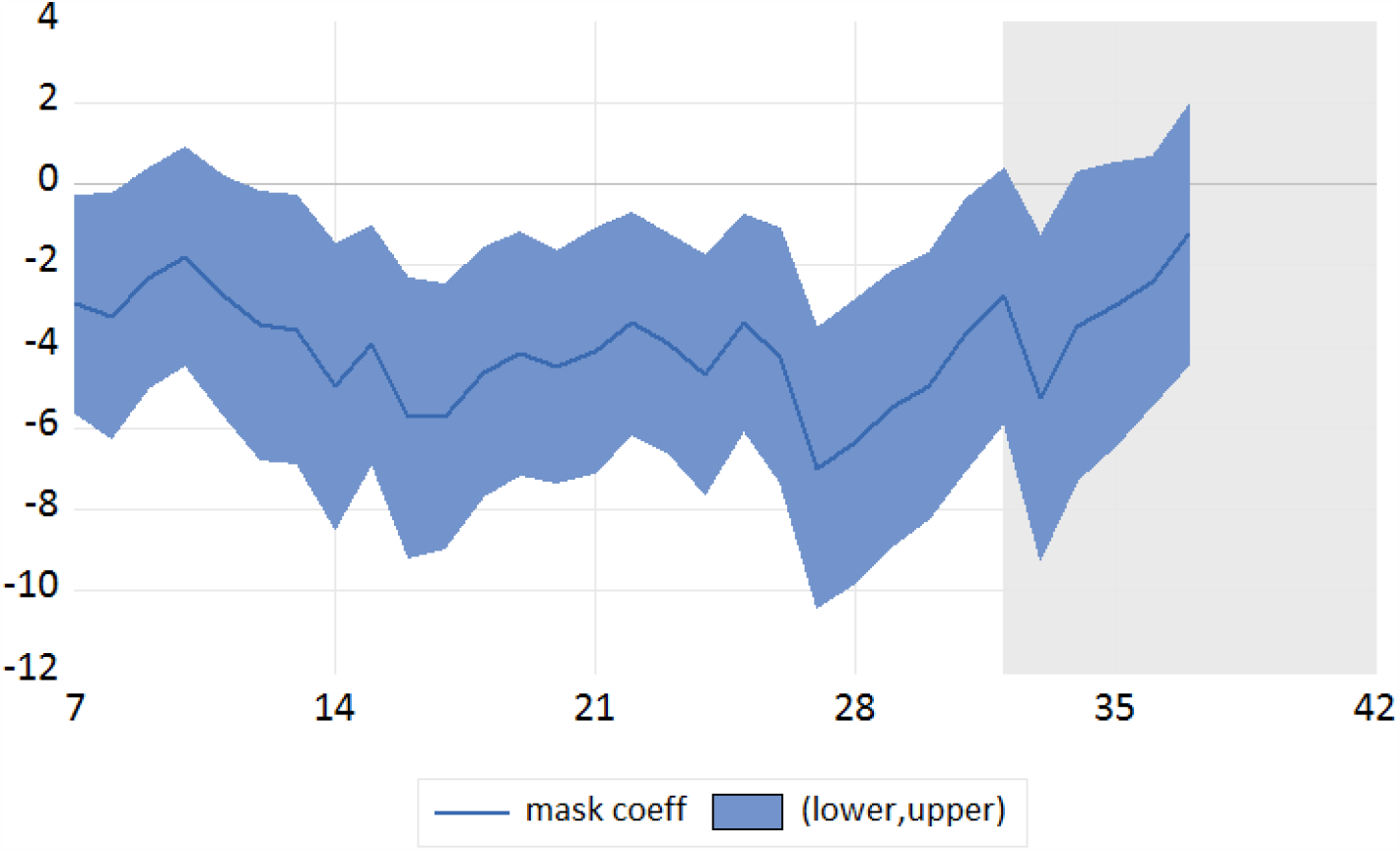
Dynamic effects of face mask use on infected cases case.

We also found a negative impact of mask wearing on fatality rates with a longer lag. Here, we consider 14- and 28-day lagged variables for illustration (Table 2). As a consequence, the mask wearing effect is more important on infections in the short term (7 to 28 days), whereas the effect is greater on fatality rates in the medium term (14 to 35 days). This is consistent with the epidemiological rationale that mask protection today can reduce the probability of fatalities after approximately one month, which is based on the dynamics of the epidemic, the incubation period, and the length of time before infection, symptoms, and complications are declared. The lag is thus greater for fatalities than for confirmation of infections.

**Table 2:** Masks effects on Covid-19 fatalities.

|  | (1)<br>deathpop | (2)<br>deathpop | (3)<br>deathpop | (4)<br>deathpop |
| --- | --- | --- | --- | --- |
| deathpop (t-1) | -0.00230<br>(0.0201) | -0.0338**<br>(0.0159) | -0.0651***<br>(0.0182) | -0.0885***<br>(0.0227) |
| deathpop (t-14) | 0.0308*<br>(0.0170) | 0.0303*<br>(0.0182) | 0.0352<br>(0.0232) | 0.0350<br>(0.0279) |
| casepop (t-14) | 0.000730<br>(0.000565) | 4.39e-06<br>(0.000719) | -0.000275<br>(0.000570) | 2.06e-05<br>(0.000625) |
| masks (t-14) | -6.47e-10*<br>(3.70e-10) |  |  |  |
| masks (t-28) |  | -1.15e-09***<br>(4.09e-10) |  |  |
| masks (t-42) |  |  | -6.23e-10<br>(6.51e-10) |  |
| masks (t-56) |  |  |  | -5.36e-10<br>(5.86e-10) |
| time | -5.69e-11<br>(4.84e-10) | 1.09e-10<br>(6.24e-10) | 8.25e-10<br>(1.18e-09) | -1.71e-09<br>(1.38e-09) |
| Constant | 1.48e-06<br>(1.07e-05) | -2.28e-06<br>(1.38e-05) | -1.81e-05<br>(2.61e-05) | 3.79e-05<br>(3.03e-05) |
| Observations | 6,687 | 5,343 | 3,999 | 2,655 |
| Number of id | 96 | 96 | 96 | 96 |

Standard errors in parentheses. $t - k$ denotes current time minus $k$ days.
\*\*\* p<0.01, \*\* p<0.05, \* p<0.1

We conducted a variety of robustness checks (SI Appendix). Considering the fact that the Covid-19 epidemic did not begin at the same time on all continents, we considered European countries for which data was available as a homogeneous and robust sample (SI Appendix, Tables S5–S11). The robustness of the sample is supported by the fact that European countries report a particularly high number of Facebook panel responses for the mask wearing variable.

We also accounted for potential omitted bias and collinearity between mask wearing and other mitigation measures, such as testing policies, school closures, and travel bans. We disentangled the effects of mask wearing from those of other mitigation measures and found a robust negative effect of mask use on the Covid-19 outbreak (Table 3), taking into account the daily number of new tests per thousand of population and the policy stringency index. The mask wearing effect appears to be robust to the introduction of other controls, such as individual mobility measures – see (27) and (28) for mobility studies – from mobile device location pings (here, we use walking and driving indexes from Apple Trend Reports), temperatures, and other mitigation measures, including Covid-19 testing policy and the Covid-19 policy stringency index.

**Table 3:** Masks effects on Covid-19 infected cases and fatalities by controling testing policy.

| VARIABLES | (1)<br>casepop | (2)<br>casepop | (3)<br>deathpop | (4)<br>deathpop |
| --- | --- | --- | --- | --- |
| casepop (t-1) | 0.161***<br>(0.0394) | 0.107***<br>(0.0284) |  |  |
| casepop (t-14) | 0.115***<br>(0.0269) | 0.117***<br>(0.0236) | 0.000802<br>(0.000710) | 0.000715<br>(0.000585) |
| masks(t-14) | -6.05e-08***<br>(2.26e-08) | -5.73e-08***<br>(1.73e-08) | -1.18e-09**<br>(5.34e-10) | -7.59e-10*<br>(4.05e-10) |
| new tests (t-14) | -1.58e-07<br>(6.55e-07) |  | 1.26e-07***<br>(4.23e-08) |  |
| stringent (t-14) |  | 9.93e-11<br>(2.21e-08) |  | -5.99e-10<br>(6.23e-10) |
| deathpop (t-1) |  |  | 0.000710<br>(0.0275) | -0.0152<br>(0.0204) |
| deathpop (t-14) |  |  | 0.0111<br>(0.0183) | 0.0281*<br>(0.0170) |
| time | 4.90e-08**<br>(1.97e-08) | 4.90e-08***<br>(1.86e-08) | -1.30e-09<br>(8.96e-10) | -4.11e-10<br>(5.45e-10) |
| Constant | -0.00107**<br>(0.000435) | -0.00108***<br>(0.000411) | 2.89e-05<br>(1.98e-05) | 9.40e-06<br>(1.21e-05) |
| Observations | 4,351 | 6,596 | 4,351 | 6,596 |
| Number of id | 68 | 95 | 68 | 95 |

Standard errors in parentheses. $t - 14$ denotes current time minus 14 days. new tests: new daily tests per thousand; \*\*\* $p < 0.01$ , \*\* $p < 0.05$ , \* $p < 0.1$

## What are the drivers of mask wearing on a global scale?

As demonstrated in the previous section, face mask use is negatively correlated with infections and fatality rates. The next question is why individuals in some countries are more likely to wear masks wearing than those in other countries. Since the main characteristics of the countries are fixed over time, it was only possible to conduct descriptive statistics investigations and cross-sectional regressions. Based on information from scatter plots, a correlation coefficients table, partial regressions (see appendix Table S2), and multivariate cross-section regressions (Tables 2 and 3), we can report some interesting insights.

First, we found that individuals appear to have implicitly understood the crucial role of mask wearing by themselves since we note a convergence to higher levels of mask wearing across countries over time. This is indicated in Figures 2 and 3 by the fact that the distribution of the mask wearing variable moves to the right as we move from the initial period (April, 23 or 24) to the final period (July, 15). In other words, more and more individuals declared on Facebook that they had adopted mask wearing to combat the Covid-19 virus. The increasing dynamics of the pandemics, the communication (both public and private) on the role of masks in reducing transmission, and the changing of habits are potential explanations for this result.

**Figure 2:**
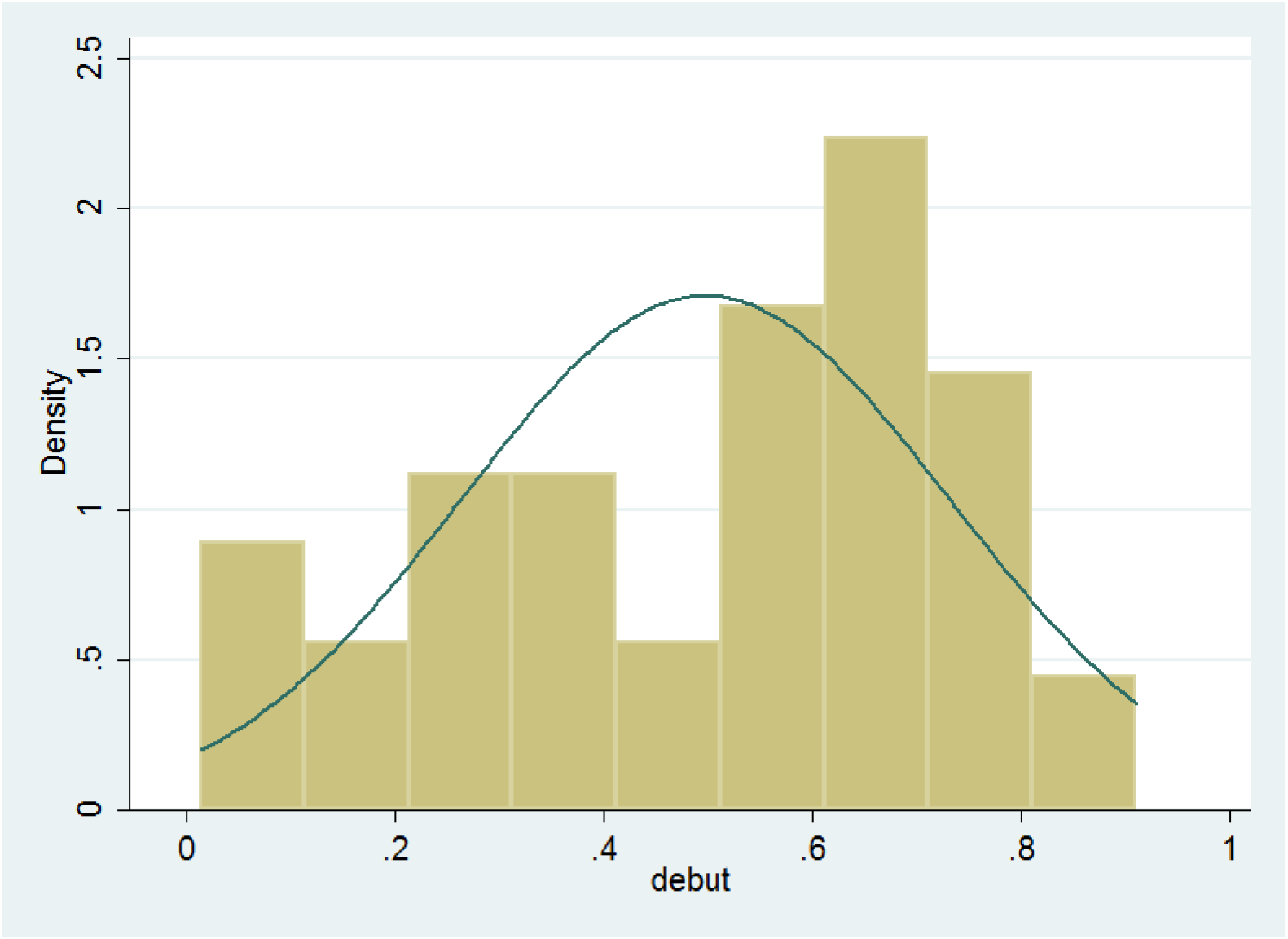
Distribution of masks wearing across countries on 23th April, 2020.

**Figure 3:**
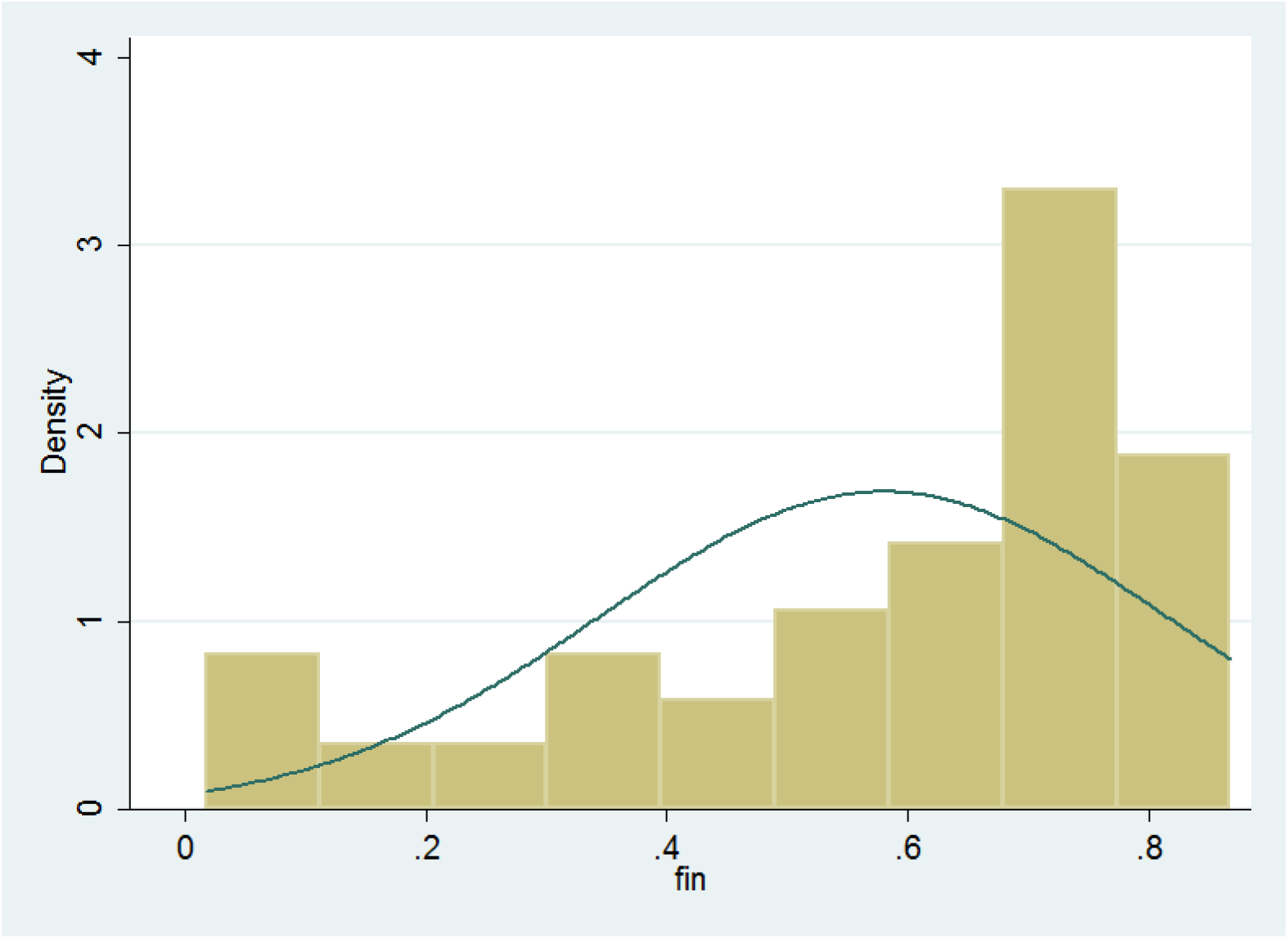
Distribution of masks wearing across countries on 15th July, 2020.

We then estimated the determinants of mask adoption on a global scale by screening a set of potential predictors (see Methods). The cross-sectional results are presented in Tables 4 and 5. We found that population density was positively associated (Fig. 4, Tables 4–5) with the percentage of individuals wearing a face mask daily. This effect is generally robust (P ¡ 0.01 to P ¡ 0.1). Mask adoption was highest in countries with a high population density and lowest in countries with a low population density. The literature shows that mask wearing is usually viewed as a complement to social distancing measures; therefore, using masks as a protective measure is more important in densely populated environments (e.g., urban areas, public transport, supermarkets, town centers) than in less densely populated regions (e.g., forests, rural areas). In state capitals and large cities, mask wearing is generally mandatory, especially on public transport. However, when we tested the urban population percentage variable directly, we found the results were less clear cut and that the associated coefficient was never significant. Population density in general seems to be more important than the urban population percentage.

**Table 4:** Cross-crountry determinants of mask covering.

| VARIABLES | (1)<br>masks | (2)<br>masks | (3)<br>masks | (4)<br>masks | (5)<br>masks |
| --- | --- | --- | --- | --- | --- |
| density | 0.0351*<br>(0.0195) | 0.0394*<br>(0.0199) | 0.0523**<br>(0.0209) | 0.0337*<br>(0.0198) | 0.0456**<br>(0.0183) |
| co2 | -0.328***<br>(0.106) | -0.300***<br>(0.102) | -0.344***<br>(0.110) | -0.319***<br>(0.101) | -0.343***<br>(0.106) |
| co2<sup>2</sup> | 0.0157***<br>(0.00498) | 0.0148***<br>(0.00492) | 0.0169***<br>(0.00524) | 0.0154***<br>(0.00483) | 0.0163***<br>(0.00510) |
| gdp |  | -0.0465*<br>(0.0271) |  |  |  |
| gov eff |  |  | -0.0839***<br>(0.0298) |  |  |
| urban |  |  |  | -0.000717<br>(0.00125) |  |
| stringency |  |  |  |  | 0.00512***<br>(0.00114) |
| Constant | 2.098***<br>(0.529) | 2.350***<br>(0.634) | 2.065***<br>(0.549) | 2.092***<br>(0.529) | 1.829***<br>(0.514) |
| Observations | 88 | 87 | 85 | 88 | 85 |
| R-squared | 0.105 | 0.142 | 0.227 | 0.108 | 0.310 |

Robust standard errors in parentheses
\*\*\* p<0.01, \*\* p<0.05, \* p<0.1

**Table 5:** Cross-crountry determinants of mask covering.

| VARIABLES | (1)<br>masks | (2)<br>masks | (3)<br>masks | (4)<br>masks |
| --- | --- | --- | --- | --- |
| density | 0.0483**<br>(0.0196) | 0.0569***<br>(0.0209) | 0.0418<br>(0.0260) | 0.0331<br>(0.0342) |
| co2 | -0.335***<br>(0.113) | -0.383***<br>(0.120) | -0.287**<br>(0.131) | -0.311<br>(0.302) |
| co2<sup>2</sup> | 0.0160***<br>(0.00555) | 0.0183***<br>(0.00579) | 0.0140**<br>(0.00630) | 0.0157<br>(0.0135) |
| stringency | 0.00468***<br>(0.00135) | 0.00418***<br>(0.00132) | 0.00623***<br>(0.00158) |  |
| diabetic | 0.00106<br>(0.00649) | 0.00232<br>(0.00687) |  |  |
| age65 | 0.000610<br>(0.00547) | 0.00515<br>(0.00520) |  |  |
| cum. cases | 0.0495<br>(0.0439) | 0.0575<br>(0.0346) | 0.0244<br>(0.0857) | 0.127*<br>(0.0749) |
| gdp | -0.0235<br>(0.0342) |  |  |  |
| gov effect |  | -0.0702*<br>(0.0358) |  |  |
| altruism |  |  | 0.0280<br>(0.0641) |  |
| tolerance |  |  |  | 0.0487**<br>(0.0204) |
| tolerance<sup>2</sup> |  |  |  | -0.000428***<br>(0.000150) |
| Constant | 1.992***<br>(0.705) | 1.957***<br>(0.599) | 1.429**<br>(0.643) | 0.574<br>(1.887) |
| Observations | 82 | 80 | 54 | 49 |
| R-squared | 0.333 | 0.362 | 0.352 | 0.485 |

Robust standard errors in parentheses
\*\*\* p<0.01, \*\* p<0.05, \* p<0.1

**Figure 4:**
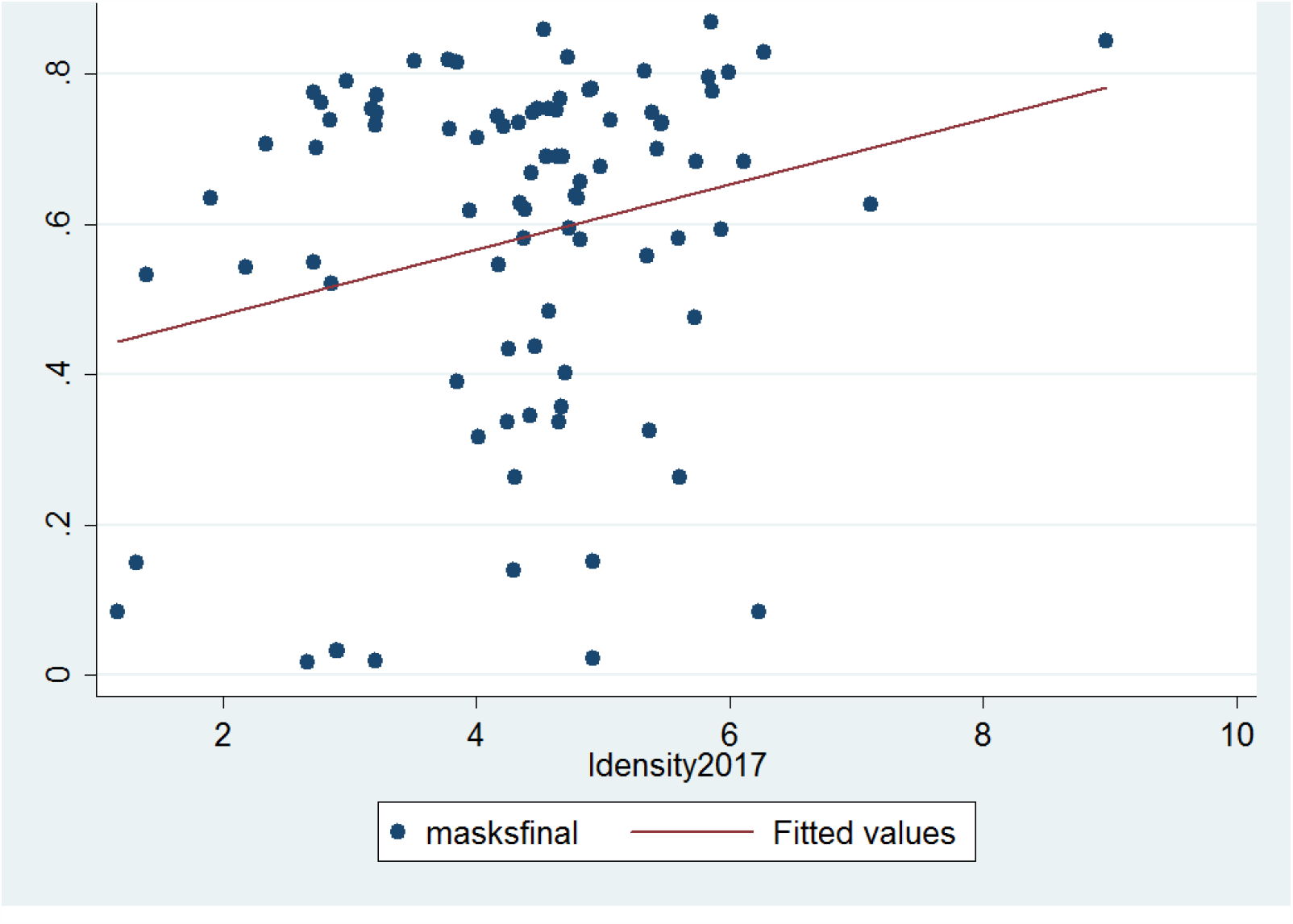
Mask adoption versus population density.

Another main driver of mask wearing on a global scale is the level of pollution. Indeed, pollution levels proxied by CO2 emissions were positively associated (P ¡ 0.01 in most cases) with the proportion of mask users only for high levels of CO2 emissions, but this effect was reversed for low levels of emissions (Fig. 5). It is probable that inhabitants of countries with high levels of pollution are more likely to wear masks to fight Covid-19 because they are in the habit of using them as a protective measure against harmful particles. Indeed, in some countries, face masks are used to reduce the negative effects of pollution in normal ‘non-Covid-19’ times. Thus, the marginal cost of adopting mask wearing behavior in those countries is low or null. No change of habits is required for individuals from these countries as they just continue to wear masks as they had done prior to the pandemic. Recent studies have also shown that mask wearing is especially necessary in highly polluted environments as Covid-19 fatality rates are exacerbated by high levels of pollution (29).

**Figure 5:**
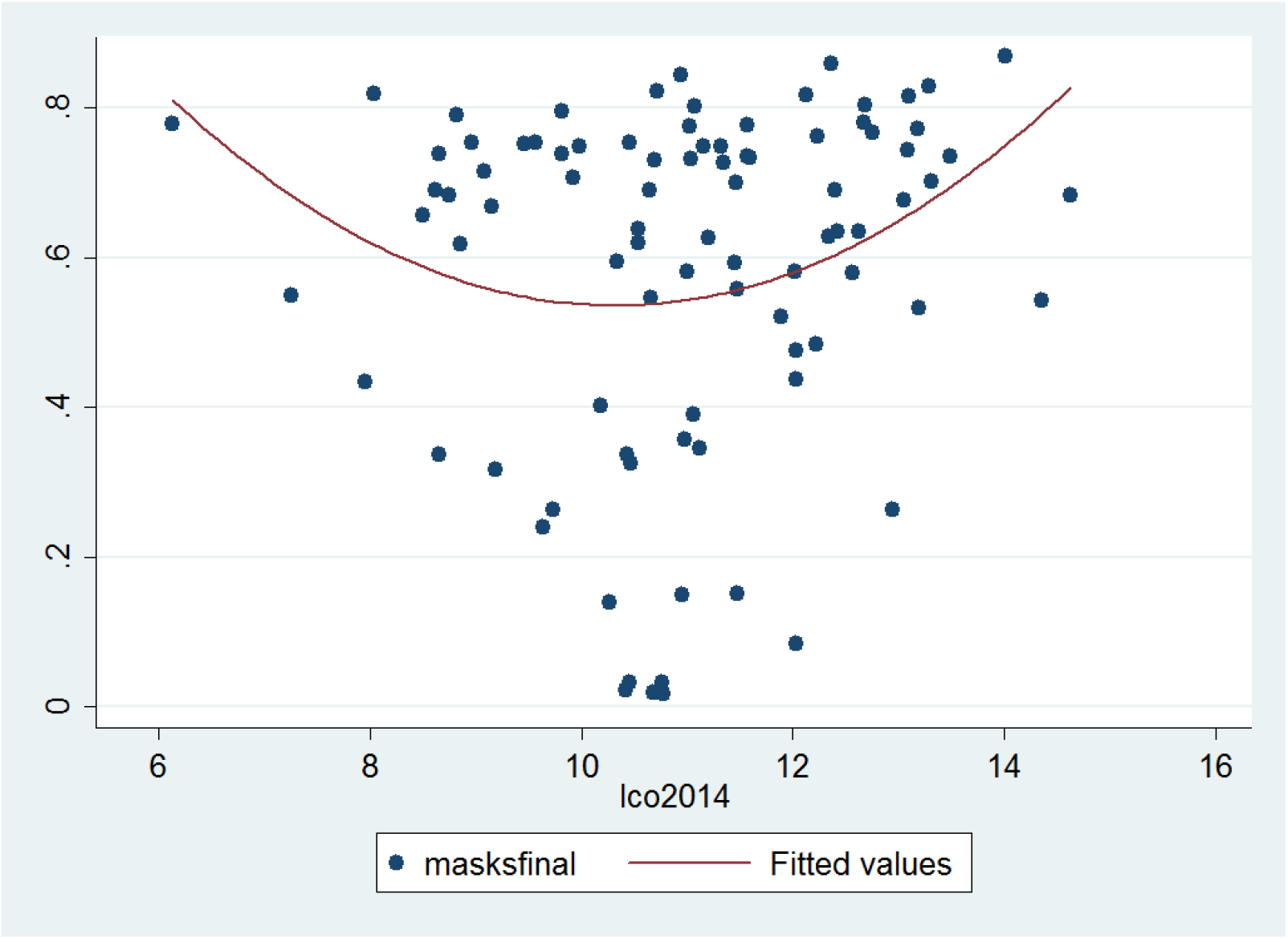
Mask adoption versus CO2 emissions.

The most important predictor of the level of mask wearing in a given population was the Covid-19 stringency index. This index, compiled by (30), measures the stringency of government responses to the Covid-19 pandemic across the world based on nine indicators: a higher score indicates a stricter government response, here on July, 15, 2020. This index was positively correlated with mask wearing, and this effect was particularly robust (P ¡ 0.001). In other words, when government responses are stricter, mask use is significantly higher (Fig. 6). If the results we report in the previous section are correct – that is, if increasing mask use reduces the negative consequences of the Covid-19 virus – the more stringent countries may obtain better results in fighting the pandemic. We also tested the lagged stringency index (one month lagged variable) and the growth rate of the index over the month previous to the date of variables counting (July, 15, 2020).

**Figure 6:**
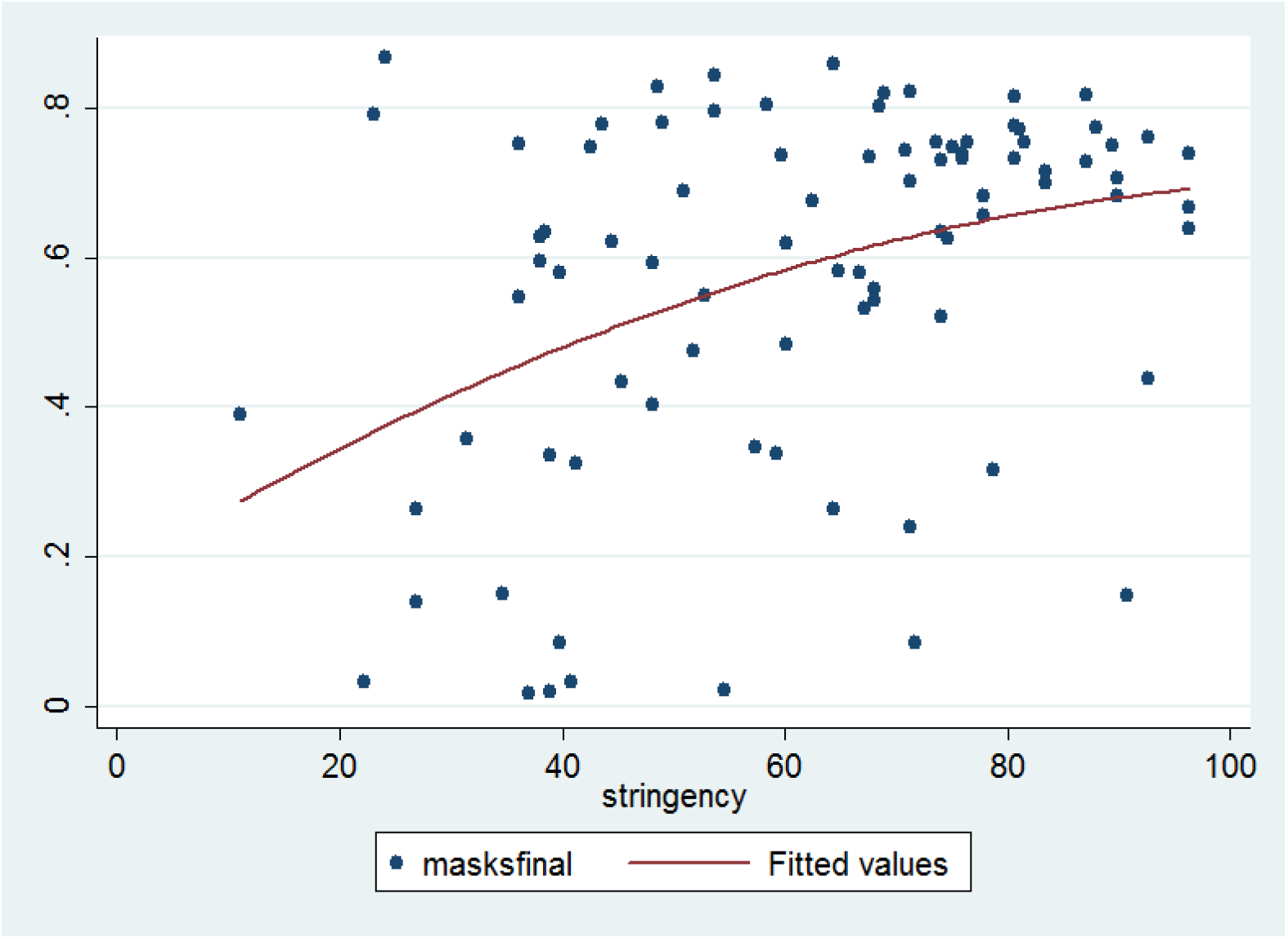
Mask adoption versus policy stringency index.

In addition, we tested whether the proportion of vulnerable individuals in a population affected the likelihood of individuals to adopt mask wearing to protect themselves and others. Linear partial regressions (SI Appendix Tables S13–S16) show that a high proportion of diabetic individuals in a population is positively associated with mask wearing. However, this effect appears not to be robust in a multivariable framework (Table 5). Similar way, results are inconclusive regarding the proportion of overweight individuals variable (SI Appendix Tables S13–S16).

Government effectiveness and GDP appear to be negatively correlated (P ¡ 0.01 in most cases) with the proportion of people wearing masks. This is surprising since we expected that rich countries would be more likely to make masks free of charge or subsidize part of the cost and that individuals in these countries would have higher incomes and, therefore, less economic constraints to prevent them from buying the required quantity of face masks to protect them in public and private areas. Regarding the government effectiveness variable, countries with a high level of government effectiveness are generally characterized by better policy formulation and implementation and higher credibility of government commitment, which is likely to increase the probability of mask wearing and higher compliance with government policy. Indeed, it has been found that strong public guidance is necessary to promote and enforce mask wearing (31 and also 25). It should be noted countries with high a GDP and those with a high government effectiveness are likely to be similar (the correlation is around 0.81 between these two variables), which explains why both coefficients are moving in the same direction. To avoid multicollinearity issues, we alternatively added government effectiveness or GDP in the tested models.

We expected that countries with a high proportion of elderly individuals are expected would have a high level of mask wearing, based on previous findings from (23). However, the sign of association between masks and the proportion of elderly individuals was negative in partial regressions (SI Appendix Tables S13–S16) and not significantly robust in multivariable regressions (Table 5). There are several possible explanations for this: countries with a high proportion of individuals aged over 65 are generally industrialized countries in Europe where, unlike in Asian countries with younger populations, mask wearing is not habitual. It is possible that older individuals, whose flexibility and capacity for adaptability are generally lower, are less likely to wear masks. Countries with a high proportion of elderly individuals are also likely to be more conservative and, thus, their inhabitants may take longer to subscribe to new habits and rules. In a letter, some authors underlined that socially responsible behaviors may not be intuitive and that behavior change takes time (32). This is probably truer for countries with older populations. Our multivariate regressions results do not indicate any significant negative relationships, which suggests that demographics structure is probably not a key factor affecting mask wearing disparities across countries.

It is also crucial to consider how the current dynamics of the pandemic might influence the habits of individuals regarding mask wearing and the adoption of other social measures to reduce the transmission rate. Public information on the dynamics and intensity of the Covid-19 epidemic is probably an important driver of mask wearing, and this variable indirectly captures this effect. When the number of cases is increasing, individuals are more informed and aware of the epidemic’s negative consequences and are, therefore, probably more inclined to take control measures such as mask wearing and social distancing more seriously. The higher the rate of infected cases, the higher the probability that individuals will personally know infected individuals (e.g., neighbors, family members, work colleagues). This may make individuals perceive the virus as more frightening and lead to increasing levels of social responsibility, making individuals more likely to wear a mask and obey rules to reduce the transmission rate. We found that the link between the infection count (we used the official count on July, 15, 2020 but also tested a lagged variable) and mask wearing was positive but not always significant at high confidence levels (Table 5).

Finally, we attempted to evaluate whether individuals in countries with higher levels of altruism, solidarity, and tolerance were more likely to use masks. A scatter plot showed a very modest positive association between mask wearing and the recent (2018) altruism index of Falk and co-authors (33). However, we found no clear-cut correlation, especially in our multivariable analysis (Table 5). We also tested the effect of the tolerance index from the World Value Survey database (34). Tolerance level was significantly associated with mask use for countries with low to intermediate tolerance values, but the association was negative for countries with high tolerance levels (SI Appendix, Table S14). This effect may be due to the fact that Scandinavian countries, which are characterized by high levels of tolerance, have implemented liberal policies on mask wearing. We also test if the trust in politicians and government can affect the mask wearing proportion in line with (5) but do not find a clear significant relationship: in other words, we do not find that a high proportion of people that do not trust in government ‘at all’ is associated to a weak proportion of mask wearing in the population but due to the limited number of observations, the inference work should be interpreted with cautious (SI Appendix Table S20). More generally, note that the number of available observations for regressions including altruism and tolerance variables is limited (around 50 observations).

Although we expected that education levels might affect the learning of useful hygiene rules and suitable behaviors to confront contagious diseases, education level (schooling variable from the World Bank or Pisa score) was found to have a non-significant negative effect on mask wearing (see Appendix Table S17).

In sum, our results complement those of previous studies, extending them from the level of individual surveys to the country scale. While previous studies investigated factors such as age, gender, or threat perception, our study explores the role of country-level macroeconomic and socio-economic determinants in explaining the substantial heterogeneity across countries regarding the wearing of face masks.

## Discussion and Conclusion

Understanding the links between face mask use and Covid-19 is crucial since the effectiveness of masks has been widely debated and contested among the general public. Our panel econometric exercise demonstrates that the wearing of face masks is negatively associated with infections and fatalities at the country level. Therefore, mask wearing has an important role to play in controlling the spread of Covid-19. Given the effectiveness of mask wearing in significantly curbing the transmission of the airborne Covid-19 virus and, thus, reducing the number of infections and fatalities, it would be helpful for governments to focus more on promoting the use of masks than on invoking the precautionary principle. We document a process of convergence over time in favor of mask adoption by showing the increasing percentages of people wearing masks across countries. This is good news as it suggests that individuals have implicitly understood the substantial impact of mask use on fighting the Covid-19 pandemic. However, our data shows that the mean proportion of inhabitants wearing masks was only 56% on July, 15, with a large standard deviation. Even in countries with high levels of mask use, the number of individuals that avoid mask wearing is probably still too large.

### Implications for policy

Appropriate public hygiene and control policies would consist of mandatory mask wearing. Given the clear effect of mask wearing on infections and fatalities and the fact that a mandatory mask policy involves little economic disruption (19) and is economically attractive, we also recommend extending mandatory face mask use even for children (over 6 years of age). Attitudes regarding altruism, tolerance, and solidarity do not appear to be sufficient to achieve the necessary levels of mask wearing. In contrast, strict government responses significantly increase levels of mask use. In this respect, our results are in line with the findings of (25), which show that a voluntary policy leads to insufficient compliance. In addition, since the absence of economic constraints that might prevent the purchase of masks in high-income countries seems not to be a key driver, the main reasons for not wearing masks appear to be subjective, cultural, or the result of a lack of legal sanctions. Legally requiring people to wear face masks, thus, appears to be an effective instrument (22). Therefore, some countries could introduce stronger penalties for not wearing masks. Indeed, the only effective way of enhancing mask adoption and saving more lives appears to be to implement more stringent policies in line with some Asian countries, such as Singapore.

### Panel data and sample issues

We conducted an original empirical work based on a 96 countries dataset between the first of January and the 15th of July 2020. For the first section, we obtain a panel with 96 countries and around 7359 observations. We collected (1) the number of confirmed COVID-19 cases and deaths for the countries in our sample from the European Center for Disease, Control and Prevention between 1st January 2020 and July, 15th 2020, the estimated population in 2019 from the World Bank’s World Development Indicators database, and the mask wearing variable from the Barkay et al. study from Maryland University using the Facebook platform (26).

Our mask wearing variable is based on the API study (26). The surveys ask respondents how many people in their household are experiencing COVID-like symptoms, among other questions. These surveys are voluntary, and individual survey responses are held by University of Maryland and are shareable with other health researchers under a data use agreement. No individual survey responses are shared back to Facebook.

Using this survey response data, we estimate the percentage of people in a given geographic region that use face mask cover. We use the smoothed weighted percentage of survey respondents that have reported use mask cover.

The descriptive statistics (average mask wearing proportion) seem coherent with other sources. For example, data based on Health Metrics and Evaluation at the University of Washington in Seattle reported in USA. We use a standardized value as a benchmark variable by dividing the mean value of wearing mask by its standard deviation. We get success to collect data for 96 countries. The number of observations varies across countries.

The dimension of our panel (96 countries) has been constrained by data availability about our main interest ‘mask’ variable. Thus, the countries in our sample have not encountered the first wave of the pandemic quite in the same time. In addition, the responsiveness rate about the mask wearing variable can be different between high-income and development countries. Considering only European countries as a robustness check (SI Appendix, Tables S5-S11) enables us to focus on countries with high responsiveness levels about the mask wearing variable to maximize the quality of the available information for this variable of great interest in our study. In addition, we have to take into account the fact that we have a panel of 96 countries with an important heterogeneity concerning the take-off Covid-19 periods (time with the first infected people) and so different Covid-19 dynamics over time: the first wave of Covid-19 epidemic has started later in Brazil than in Italy. Considering only homogeneous European countries is therefore a mean to test the presence of sample bias. Since the number of observations varies across countries, we conduct a robustness check by only considering European countries (SI Appendix, Tables S5-S11). Results reported on 21 available European countries in our database are completely in accordance with the global results conducted on 96 countries.

### Dynamic econometric panel model to assess mask effectiveness

To test our theoretical assumptions, we use a dynamic panel econometric model as follows:

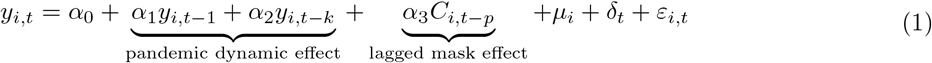

Where the subscripts *i* and *t* represent country index and periods (days) respectively. The dependent variable, *y*_*i,t*_, can be the number of infected individuals (casespop) or deaths (deathpop) per capita (considering the population size) at time *t. C*_*i,t*−*p*_ is a vector of variables depicting the effects of meteorological conditions in day *t* − *p*. Country-specific fixed effect, *µ*_*i*_, are included to control for time-invariant omitted-variable bias and *ε*_*i,t*_ is the error term. *δ*_*t*_ is a deterministic time trend that controls the deterministic dynamics of the epidemics over the studied period and captures some unobserved information about the pandemic common to all countries. In addition, lagged terms *y*_*i,t*−*k*_ capture the stochastic part of the pandemic dynamics. Indeed, considering lags in epidemic dynamics is crucial for identification concerns. In addition, the literature (6 and 21) have identified that threat perceptions about the current crisis can change the mask adoption. We assume *k* equal to 7 or 14 lags/days in our baselines specifications considering incubation and confirmation periods (see for instance (36) about the incubation period) presented in the Covid-19 literature. Moreover, for logical reasons, since the mask wearing do not immediately impact the Covid-19 spread, the mask wearing variables are also included in our model with a lag of order *p*. Indeed, there are delays between the time of potential avoided infection corresponding to mask wearing and the time of official counting of a potential infected individuals (or fatality). Therefore, when dealing with *p*, a minimum of 7 to 14 days is considered. A benchmark 28 lags (about one month) delay is considered when dealing with *deathpop* because of the more important lag lenght assumed between the infection and the deaths related to the Covid-19 virus. More longer delays of 42 and 56 days have been also considered in robustness checks to take into account the dynamic persistence of the pandemic (see also SI Appendix) and the delay between the infection time and potential health problems. *casepop* and *deathpop* respectively give a short-run and medium/long-run time perspective of the dramatic outcomes of the pandemic. Note that when *y*_*i,t*_ is the fatality rate, we also add the ratio of infected cases per capita in our benchmark specification in order to account for the fact that the level of the pandemic can impact the fatality rate. The reason behind is to control for a level effect and a kind of saturation effect of the health system (too many infected individuals to manage is likely to finally increase the fatality rate).

### Identification issues

Equation (1) can be estimated by the mean group (MG) estimator introduced by (37). Both estimators are relevant for macro panels such as the one used in this paper: *T* is equal to 84 and is thus close to *N* = 96 (see SI Appendix). We also used the dynamic fixed effects (DFE) estimates and many robustness regressions and tests to validate the consistency of our results (SI Appendix, Tables S10-S11). The Mean Group (MG) estimator consists in estimating each regression separately for each panel member *i* (country here) with a minimum of restrictions. All estimated coefficients are heterogeneous and are subsequently averaged across countries *via* a simple unweighted average (37-39). An intercept is included to capture country fixed effects as well as a linear trend. In the dynamic fixed effects (DFE), the slopes are homogeneous but the intercepts are allowed to vary across countries (37-39).

Although we apply appropriate macro-panel estimators to our data, several issues can nonetheless emerge. First, using dynamic models, we are vulnerable to the so-called Nickel bias. Here, this bias is relatively negligible, notably considering the important time length of our series. Second, panel regressions may be exposed to an omitted variables bias. It would be possible to include control variables such as other control measures (e.g. testing, travel controls) or structural determinants (e.g. population density and demographics such as the population over 65, tourists flows, GDP per capita, and measures of health infrastructures). Considering the so-called problem of ‘bad controls’, our set of explanatory variables is assumed to be restricted to the mask variable in order to avoid an over-controlling problem (40). In addition, considering data availability and the fact they are time-invariant variables, we capture these unobservables *via* the lagged term *y*_*i,t*−1_ and above all with country fixed effects. Another identification issue is related to the potential reverse causality bias related to our Covid-19 variables: news about contemporaneous dynamics of the Covid-19 outbreaks and counts can change the human behavior in real time and the social distancing. This is why lags of dependent variables must be added in our model. Finally, persistence and multicollinearity are other usual issues in panel studies. We have controlled for both by computing autocorrelations LM (Lagrange Multiplier) tests and VIF/Tolerance ratios after each estimated regression. In SI Appendix, we also consider endogeneity issues and other tests related to the specification of our econometric model, the choice of an alternative estimator, and several changes in the sample composition.

### Cross-section model to investigate face mask adoption

We collect data from different sources (SI Appendix) to estimate a very simple cross-section model of the following form at a country level:

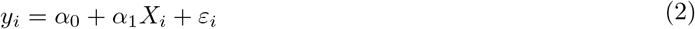

where *y*_*i*_ captures proportion of individuals declaring wearing a face mask on 15th of July, 2020 and *X*_*i*_ is a set of control variables including population density, CO2 emissions (and squared CO2 emissions), GDP level (in logarithms), government effectiveness index, altruism index, tolerance index, Covid-19 policy stringency index, urban population percentage, education level, diabetic population percentage and overweight population percentage have been considered. We also assume that the dynamics of the epidemics proxied by the count of cases is also a potential driver of masks wearing propensity. See SI Appendix for complete information about definitions and sources of the data. We also test as a robustness check the same model with mask wearing data on April, 23, 2020 corresponding to the first available and oldest data from the Maryland survey (SI Appendix). Other variables (overweight part of the population, testing, surveillance, travel restrictions and school closures policies) have been introduced (SI Appendix, Tables S18-S19).

## Data Availability

The data will be available on a public website when the paper will be published by an academic journal but are available upon request (by email).

https://sites.google.com/site/olivierdamette/research

## Author Affiliations

### Data Archival

SI dataset in .dat format and codes are available upon request.

## Supporting Information Appendix (SI)

SI Appendix is available at https://sites.google.com/site/olivierdamette/research

