## Supplementary material for "Zorro versus Covid-19: fighting the pandemic with face masks": SI Appendix

December 14, 2020

### 1 Contents

In this SI Appendix, we give an extensive information about statistical analysis, variables definition and sources and selected countries. We also outline lot of robustness checks by extending the results from Tables 1-5 of the paper by supplementary specifications. The maximum number of observations available is around 7359.

### 2 Effectiveness of mask use: additional regressions and robustness checks

We build a panel database of 96 available countries between 1st January, 2020 and July, 15, 2020. We obtain an unbalanced panel data set with a daily frequency.

#### 2.1 Taking into account additional control policies variables

The existence of a significant correlation between the number of mas wearing people and the number of infected cases and fatalities respectively is a controversial special issue. Some critiques have been addressed to Zhang et al. (16), especially the fact that they do not consider other non pharmaceutical mitigation policies in their statistical analysis as well as the length of the studied sample, notably in the New-York case.

Here, we consider additional control variables and especially include variables about other non pharmaceutical mitigation policies. Considering our time varying panel day by day database, we should incorporate some time varying variables to proxy other policy measures. We first consider the number of new tests per 100k inhabitants to investigate if the mask wearing effect is not driven by the existence of a simultaneous testing policy. We find that the effect of mask wearing is robust to the presence of a 'tests' variable for 14 days in the paper (Table 3) but we extend this test to more lags (Tables 1 and 2 from this appendix). As a consequence, the mask effect is not falsely significant due to the potential success of a simultaneous testing control policy.

The Tables 1 and 2 display the results for our global sample for infected cases and fatalities respectively (as ratios). Due to missing observations for some countries for the *new_tests* variable, the total number of observations is comprised between 3500 and 4803 for 67 available countries (over 96 initially).

We also add the stringency index (*stringency*) in Tables 3 and 4 to control the existence of other control/lockdown measures in the same time the masks have been used. Thus, we are able to disentangle the effects from the masks and the effects stemming from other control and mitigation measures. The mask use effect is negative and significant at high significance level (P¡0.01) for lags 14 and 28 (infected cases) and lag 14 and 42 for fatality rates regressions (at a lower significance level).

Table 1: Fatality rate model with tests per 100k control variable as an additional control, global sample, infected cases, MG estimates

| VARIABLES | (1)<br>casepop | (2)<br>casepop | (3)<br>casepop | (4)<br>casepop |
|---|---|---|---|---|
| casepop (t-1) | 0.179*** | 0.161*** | 0.121*** | 0.0285 |
| | (0.0396) | (0.0394) | (0.0396) | (0.0411) |
| casepop (t-14) | 0.105*** | 0.115*** | 0.137*** | 0.184*** |
| | (0.0255) | (0.0269) | (0.0348) | (0.0403) |
| masks (t-7) | -1.85e-08 | | | |
| | (1.52e-08) | | | |
| new tests (t-7)) | 1.60e-06 | | | |
| | (9.76e-07) | | | |
| masks(t-14) | | -6.05e-08*** | | |
| | | (2.26e-08) | | |
| new tests (t-14) | | -1.58e-07 | | |
| | | (6.55e-07) | | |
| masks (t-28) | | | -5.98e-08*** | |
| | | | (2.21e-08) | |
| new tests (t-28) | | | 1.40e-06 | |
| | | | (1.33e-06) | |
| masks (t-42) | | | | -6.66e-09 |
| | | | | (1.82e-08) |
| new tests (t-42) | | | | 1.06e-06 |
| | | | | (1.41e-06) |
| time | 4.20e-08* | 4.90e-08** | 7.90e-08*** | 8.37e-08** |
| | (2.44e-08) | (1.97e-08) | (2.76e-08) | (3.59e-08) |
| Constant | -0.000920* | -0.00107** | -0.00174*** | -0.00183** |
| | (0.000538) | (0.000435) | (0.000609) | (0.000794) |
| Observations | 4,779 | 4,351 | 3,454 | 2,558 |
| Number of id | 68 | 68 | 67 | 66 |

Standard errors in parentheses
*** p<0.01, ** p<0.05, * p<0.1

Table 2: Fatality rate model with tests per 100k control variable as an additional control, global sample, infected cases, MG estimates

| | (1) | (2) | (3) |
|---|---|---|---|
| VARIABLES | deathpop | deathpop | deathpop |
| deathpop (t-1) | 0.000710 | -0.0461** | -0.0711*** |
| | (0.0275) | (0.0210) | (0.0252) |
| deathpop (t-14) | 0.0111 | 0.0181 | 0.0485** |
| | (0.0183) | (0.0186) | (0.0226) |
| casepop (t-14) | 0.000802 | -0.000619 | -0.00125 |
| | (0.000710) | (0.000838) | (0.000891) |
| masks (t-14) | -1.18e-09** | | |
| | (5.34e-10) | | |
| new tests (t-14) | 1.26e-07*** | | |
| | (4.23e-08) | | |
| masks (t-28) | | -1.08e-09* | |
| | | (5.88e-10) | |
| new tests (t-28) | | 3.83e-08 | |
| | | (3.98e-08) | |
| masks (t-42) | | | -3.12e-10 |
| | | | (5.54e-10) |
| new tests (t-42) | | | 2.98e-08 |
| | | | (4.61e-08) |
| time | -1.30e-09 | -2.01e-10 | 2.71e-09 |
| | (8.96e-10) | (1.07e-09) | (1.91e-09) |
| Constant | 2.89e-05 | 4.56e-06 | -5.97e-05 |
| | (1.98e-05) | (2.37e-05) | (4.21e-05) |
| Observations | 4,351 | 3,454 | 2,558 |
| Number of id | 68 | 67 | 66 |

Standard errors in parentheses
*** p<0.01, ** p<0.05, * p<0.1

Table 3: Fatality rate model with tests per 100k control variable as an additional control, global sample, infected cases, MG estimates

| | (1) | (2) | (3) | (4) |
|---|---|---|---|---|
| VARIABLES | casepop | casepop | casepop | casepop |
| casepop (t-1) | 0.116*** | 0.107*** | 0.0575** | 0.00283 |
| | (0.0279) | (0.0284) | (0.0253) | (0.0277) |
| casepop (t-14) | 0.104*** | 0.117*** | 0.105*** | 0.138*** |
| | (0.0223) | (0.0236) | (0.0260) | (0.0316) |
| masks (t-7) | -1.57e-08 | | | |
| | (1.43e-08) | | | |
| stringent (t-7) | 7.85e-09 | | | |
| | (1.97e-08) | | | |
| masks (t-14) | | -5.73e-08*** | | |
| | | (1.73e-08) | | |
| stringent (t-14) | | 9.93e-11 | | |
| | | (2.21e-08) | | |
| masks (t-28) | | | -4.93e-08*** | |
| | | | (1.68e-08) | |
| stringent (t-28) | | | 1.81e-08 | |
| | | | (2.82e-08) | |
| masks (t-42) | | | | -9.00e-09 |
| | | | | (1.23e-08) |
| stringent (t-42) | | | | 1.40e-08 |
| | | | | (2.63e-08) |
| time | 4.41e-08** | 4.90e-08*** | 7.05e-08** | 8.12e-08* |
| | (1.82e-08) | (1.86e-08) | (2.80e-08) | (4.25e-08) |
| Constant | -0.000999** | -0.00108*** | -0.00155** | -0.00183* |
| | (0.000405) | (0.000411) | (0.000621) | (0.000934) |
| Observations | 7,254 | 6,596 | 5,280 | 3,959 |
| Number of id | 95 | 95 | 95 | 95 |

Standard errors in parentheses
*** p<0.01, ** p<0.05, * p<0.1

Table 4: Fatality rate model with tests per 100k control variable as an additional control, global sample, infected cases, MG estimates

| VARIABLES | (1)<br>deathpop | (2)<br>deathpop | (3)<br>deathpop | (4)<br>deathpop |
|---|---|---|---|---|
| deathpop (t-1) | -0.0251 | -0.0152 | -0.0409*** | -0.0900*** |
| | (0.0200) | (0.0204) | (0.0153) | (0.0179) |
| deathpop (t-14) | 0.0173 | 0.0281* | 0.0307* | 0.0362 |
| | (0.0183) | (0.0170) | (0.0177) | (0.0233) |
| casepop (t-14) | 0.000749 | 0.000715 | -0.000390 | -0.000848 |
| | (0.000785) | (0.000585) | (0.000834) | (0.000521) |
| masks (t-7) | -8.67e-10* | | | |
| | (4.58e-10) | | | |
| stringent (t-7) | -5.37e-10 | | | |
| | (8.90e-10) | | | |
| masks (t-14) | | -7.59e-10* | | |
| | | (4.05e-10) | | |
| stringent (t-14) | | -5.99e-10 | | |
| | | (6.23e-10) | | |
| masks (t-14) | | | -5.58e-10 | |
| | | | (4.05e-10) | |
| stringent (t-28) | | | 1.14e-09 | |
| | | | (7.65e-10) | |
| masks (t-42) | | | | -1.58e-09** |
| | | | | (7.43e-10) |
| stringent (t-42) | | | | -1.23e-09 |
| | | | | (1.04e-09) |
| time | -1.58e-10 | -4.11e-10 | 0 | -4.08e-10 |
| | (6.98e-10) | (5.45e-10) | (5.85e-10) | (1.19e-09) |
| Constant | 3.90e-06 | 9.40e-06 | -8.60e-07 | 1.12e-05 |
| | (1.54e-05) | (1.21e-05) | (1.33e-05) | (2.59e-05) |
| Observations | 7,254 | 6,596 | 5,280 | 3,959 |
| Number of id | 95 | 95 | 95 | 95 |

Standard errors in parentheses
*** p<0.01, ** p<0.05, * p<0.1

#### 2.2 Taking into account additional controls: individual mobility and temperatures

We also add a mobility 'driving' variable to investigate the impact of individual mobility (see data section for information about these Apple data). If individuals are more mobile, they probably can use more masks to go outside (at work, at school or university, at the supermarket). Above all, the mobility can be viewed as a another proxy of the control policies (lockdown, stay-at-home requirements, restrictions on public gatherings etc): if the mobility is strongly reduced, it is an other indirect indicator that the level of stringency of the control policies is high.

Considering our data about mobility and temperatures, we are working on a shorter sample of European countries. When we add other control variables, the number of countries dropped to 22 (and even 17 in one model). Before performing estimates with additional explanatory variables on this shorter sample, we check if the effect of mask wearing is robust when this sample is considered as a benchmark. For transparency concerns, we first present the results of the mask wearing impact (Table 5) before introducing additional controls: temperatures, mobility and new tests per 100k inhabitants. The mask variable is negatively associated to *casepop* with very high level of significance (P¡0.01) for 7 and 14 days/lags. Then, we add other controls in this benchmark regression (Tables 6-7).

#### 2.3 Sample bias: considering only European countries

We consider the case of European countries only, over the maximum time period (1st January to 15, July, 2020) using our benchmark MG estimator. Considering only European countries enables us to focus on countries with high responsiveness levels about the mask wearing variable maximizing the quality of the available information for this variable of great interest in our study. In addition, we have to take into account the fact that we have a panel of 96 countries with an important heterogeneity concerning the take-off Covid-19 periods (time with the first infected people) and so different Covid-19 dynamics over time: the first wave of Covid-19 epidemic has started later in Brazil than in Italy. Considering only homogeneous European countries is therefore a mean to test the presence of sample bias. Finally, we used two different estimators: MG (Mean Group) estimator and DFE (Dynamic Fixed Effects) estimator.

We consider the case of European countries over the maximum time period but using an alernative DFE (Dynamic Fixed effect) estimator instead of the MG one that has been used as a benchmark. The standardized mask variable has a significant (P¡0.1) effect on infected cases considering 14 lags and considering 7 and 14 lags for fatality rates. The maximum effect is observed for 7-14 days delay in the way of epidemiological investigations.

Table 5: Benchmark regression with a shorter sample of European countries before introducing additional controls, MG estimates

| VARIABLES | (1) casepop | (2) casepop | (3) casepop |
|---|---|---|---|
| casepop (t-1) | 0.0460 | 0.0152 | -0.199* |
| | (0.0628) | (0.0496) | (0.109) |
| casepop (t-14) | 0.0497 | 0.0484 | 0.136* |
| | (0.0326) | (0.0424) | (0.0715) |
| masks (t-7) | -0.00932*** | | |
| | (0.00289) | | |
| masks (t-14) | | -0.00633*** | |
| | | (0.00234) | |
| masks (t-28) | | | -0.00307 |
| | | | (0.00225) |
| Constant | 1.664*** | 1.452*** | 0.816*** |
| | (0.300) | (0.290) | (0.204) |
| Observations | 616 | 462 | 154 |
| Number of id_country | 22 | 22 | 22 |

Standard errors in parentheses
*** p<0.01, ** p<0.05, * p<0.1

Table 6: Infected cases with tests per 100k control variable (lagged 7 days), European countries, shorter sample, MG estimates

| | (1) | (2) |
|---|---|---|
| VARIABLES | casepop | casepop |
| casepop (t-1) | 0.00714 | -0.270* |
| | (0.0592) | (0.144) |
| casepop (t-14) | 0.0416 | 0.172* |
| | (0.0339) | (0.0982) |
| masks (t-7) | -0.00955** | |
| | (0.00410) | |
| new tests (t-7) | -0.000127 | |
| | (0.000133) | |
| masks (t-28) | | -0.00262 |
| | | (0.00944) |
| new tests (t-28) | | -0.000207 |
| | | (0.000686) |
| Constant | 1.423*** | 1.165 |
| | (0.317) | (0.833) |
| Observations | 616 | 154 |
| Number of id_country | 22 | 22 |

Standard errors in parentheses
*** p<0.01, ** p<0.05, * p<0.1

Table 7: Infected cases with temperatures and mobility/driving variables, MG estimates

| VARIABLES | (1)<br>casepop | (2)<br>casepop | (3)<br>casepop |
|---|---|---|---|
| casepop (t-1) | -0.0260 | -0.0524 | -0.0701 |
| | (0.0584) | (0.0597) | (0.0584) |
| casepop (t-14) | 0.0400 | 0.0181 | 0.00961 |
| | (0.0291) | (0.0324) | (0.0276) |
| masks (t-7) | -0.0106** | -0.0101* | -0.0103 |
| | (0.00534) | (0.00566) | (0.00712) |
| tests (t-7) | -8.60e-05 | -3.92e-05 | -5.16e-05 |
| | (0.000170) | (0.000259) | (0.000280) |
| temperature(t-7) | -0.0242 | -0.00226 | 0.00911 |
| | (0.102) | (0.153) | (0.156) |
| mobility(t-7) | | -0.00457 | -0.00518 |
| | | (0.00310) | (0.00939) |
| mobility*mask (t-7) | | | -0.000106 |
| | | | (0.000135) |
| Constant | 1.401*** | 1.125* | 1.275 |
| | (0.379) | (0.597) | (0.819) |
| Observations | 616 | 572 | 572 |
| Number of id_country | 22 | 22 | 22 |

Standard errors in parentheses. Mobility is 'driving' from Google Mobility reports.
*** p<0.01, ** p<0.05, * p<0.1

Table 8: Infected cases for European countries, MG estimates

| | (1) | (2) | (3) | (4) |
|---|---|---|---|---|
| VARIABLES | casepop | casepop | casepop | casepop |
| casepop (t-1) | 0.122** | 0.244*** | 0.226*** | 0.188*** |
| | (0.0511) | (0.0218) | (0.0219) | (0.0235) |
| casepop (t-14) | 0.136*** | 0.257*** | 0.224*** | 0.197*** |
| | (0.0353) | (0.0153) | (0.0100) | (0.00849) |
| masks (t-7) | -5.04e-08*** | | | |
| | (1.50e-08) | | | |
| masks (t-14) | | -1.12e-08* | | |
| | | (6.29e-09) | | |
| masks (t-28) | | | -2.26e-09 | |
| | | | (9.14e-09) | |
| masks (t-42) | | | | 2.72e-08 |
| | | | | (3.11e-08) |
| time | 2.01e-08 | 4.02e-08* | 4.51e-08 | -6.69e-08 |
| | (3.21e-08) | (2.29e-08) | (4.31e-08) | (1.61e-07) |
| Constant | -0.000433 | -0.000882* | -0.000989 | 0.00148 |
| | (0.000707) | (0.000505) | (0.000951) | (0.00355) |
| Observations | 1,617 | 1,470 | 1,176 | 882 |
| R-squared | | 0.183 | 0.113 | 0.071 |
| Number of id | 21 | 21 | 21 | 21 |

Standard errors in parentheses
*** p<0.01, ** p<0.05, * p<0.1

Table 9: Infected cases for European countries, MG estimates

| VARIABLES | (1) deathpop | (2) deathpop | (3) deathpop | (4) deathpop |
|---|---|---|---|---|
| deathpop (t-1) | -0.0265 | 0.0133 | 0.0173 | -0.0470 |
| | (0.0430) | (0.0407) | (0.0279) | (0.0374) |
| deathpop (t-14) | 0.0741* | 0.0627* | 0.0681** | 0.1000** |
| | (0.0425) | (0.0376) | (0.0322) | (0.0421) |
| casepop (t-14) | 0.00246 | 0.00289** | -0.000542 | -0.00241* |
| | (0.00238) | (0.00136) | (0.00177) | (0.00136) |
| masks (t-7) | -4.82e-09*** | | | |
| | (1.65e-09) | | | |
| masks (t-14) | | -4.01e-09*** | | |
| | | (1.37e-09) | | |
| masks (t-28) | | | -1.70e-09** | |
| | | | (8.55e-10) | |
| masks (t-42) | | | | -1.94e-09 |
| | | | | (1.41e-09) |
| time | -6.58e-09** | -3.92e-09** | -3.29e-09** | -1.53e-09 |
| | (2.58e-09) | (1.68e-09) | (1.55e-09) | (1.95e-09) |
| Constant | 0.000146** | 8.60e-05** | 7.37e-05** | 3.41e-05 |
| | (5.71e-05) | (3.75e-05) | (3.43e-05) | (4.29e-05) |
| Observations | 1,617 | 1,470 | 1,176 | 882 |
| Number of id | 21 | 21 | 21 | 21 |

Standard errors in parentheses

*** $p<0.01$, ** $p<0.05$, * $p<0.1$

Table 10: Infected cases for European countries, DFE estimates

| | (1) | (2) | (3) | (4) |
|---|---|---|---|---|
| VARIABLES | casepop | casepop | casepop | casepop |
| casepop (t-1) | 0.256*** | 0.244*** | 0.226*** | 0.188*** |
| | (0.0252) | (0.0218) | (0.0219) | (0.0235) |
| casepop (t-14) | 0.266*** | 0.257*** | 0.224*** | 0.197*** |
| | (0.00709) | (0.0153) | (0.0100) | (0.00849) |
| masks (t-7) | -6.66e-09 | | | |
| | (6.91e-09) | | | |
| masks (t-14) | | -1.12e-08* | | |
| | | (6.29e-09) | | |
| masks (t-28) | | | -2.26e-09 | |
| | | | (9.14e-09) | |
| masks (t-42) | | | | 2.72e-08 |
| | | | | (3.11e-08) |
| Constant | -0.000807* | -0.000882* | -0.000989 | 0.00148 |
| | (0.000425) | (0.000505) | (0.000951) | (0.00355) |
| time | 3.68e-08* | 4.02e-08* | 4.51e-08 | -6.69e-08 |
| | (1.93e-08) | (2.29e-08) | (4.31e-08) | (1.61e-07) |
| Observations | 1,617 | 1,470 | 1,176 | 882 |
| R-squared | 0.243 | 0.183 | 0.113 | 0.071 |
| Number of id | 21 | 21 | 21 | 21 |

Robust standard errors in parentheses

*** p<0.01, ** p<0.05, * p<0.1

Table 11: Fatality rates for European countries, DFE estimates

| VARIABLES | (1) deathpop | (2) deathpop | (3) deathpop | (4) deathpop | (5) deathpop |
|---|---|---|---|---|---|
| deathpop (t-1) | 0.161*** | 0.150** | 0.0879*** | 0.0379 | 0.00858 |
| | (0.0486) | (0.0558) | (0.0303) | (0.0455) | (0.0687) |
| deathpop (t-14) | 0.172* | 0.138 | 0.103 | 0.0446 | 0.0626 |
| | (0.0858) | (0.0928) | (0.129) | (0.0798) | (0.0984) |
| casepop (t-14) | 0.00825 | 0.00725 | 0.00423* | 0.00725*** | 0.00888*** |
| | (0.00710) | (0.00580) | (0.00206) | (0.000876) | (0.00209) |
| masks (t-7) | -3.39e-09* | | | | |
| | (1.84e-09) | | | | |
| masks (t-14) | | -3.75e-09* | | | |
| | | (2.09e-09) | | | |
| masks (t-28) | | | -4.33e-09 | | |
| | | | (3.85e-09) | | |
| masks (t-42) | | | | 5.23e-10 | |
| | | | | (1.59e-09) | |
| masks (t-56) | | | | | -2.46e-09 |
| | | | | | (3.88e-09) |
| time | -1.10e-08** | -1.06e-08** | -8.57e-09** | -9.82e-09* | -7.11e-09 |
| | (4.58e-09) | (4.56e-09) | (3.74e-09) | (5.58e-09) | (9.09e-09) |
| Constant | 0.000245** | 0.000234** | 0.000190** | 0.000217* | 0.000158 |
| | (0.000101) | (0.000101) | (8.29e-05) | (0.000123) | (0.000201) |
| Observations | 1,617 | 1,470 | 1,176 | 882 | 588 |
| R-squared | 0.289 | 0.187 | 0.072 | 0.034 | 0.040 |
| Number of id | 21 | 21 | 21 | 21 | 21 |

Robust standard errors in parentheses
*** p<0.01, ** p<0.05, * p<0.1

### 3 Determinants of mask use: additional statistical analysis

#### 3.1 Matrix of correlation and potential collinearity

We have checked the complete matrix of correlations to avoid collinearity problems. For example, rich countries characterized by higher GDP levels (in natural logarithms units) have a high probability to have also a high proportion of elder people and high government effectiveness index. As a consequence, a mutlivariable regression with $GDP$, $Age65$ and $goveffect$ simultaneously is not robust due to the presence of collinearity issues.

Table 12: Cross-correlation table

| Variables | density | lco2014 | stringency | lgdp17 | age652017 | gov_effe | cum cases | altruism | reg, robust |
|---|---|---|---|---|---|---|---|---|---|
| density2017 | 1.000 | | | | | | | | |
| co2 | 0.094 | 1.000 | | | | | | | |
| stringency | -0.140 | 0.029 | 1.000 | | | | | | |
| gdp | 0.116 | 0.478 | -0.347 | 1.000 | | | | | |
| age652017 | 0.082 | 0.326 | -0.597 | 0.675 | 1.000 | | | | |
| gov_effectiveness_2018 | 0.174 | 0.334 | -0.500 | 0.835 | 0.715 | 1.000 | | | |
| cum cases | 0.110 | 0.148 | 0.217 | 0.424 | -0.038 | 0.234 | 1.000 | | |
| altruism | 0.048 | -0.195 | 0.136 | -0.287 | -0.252 | -0.182 | -0.041 | 1.000 | |
| reg , robust | -0.168 | -0.150 | -0.087 | 0.415 | 0.283 | 0.475 | 0.062 | -0.010 | 1.000 |

#### 3.2 Partial linear regressions

In our cross-sectional database, we select mask use proportion variable on July, 15, 2020 corresponding to the latest observation of our daily frequency panel dataset. We also select the mask use proportion variable in the beginning period (this date varies from April, 23 to April, 25, 2020 according to the countries) to take into account the dynamics of the pandemic and the potential adjustments in human behaviors regarding the mask wearing as well as the effects of the control policies implemented by the governments.

We have tested partial linear regressions for all potential determinants. We have considered the presence of potential quadratic forms. In the following table X, we present the most suitable specifications and only report the most significant regression: for instance, population density regression does not incorporate significant quadratic variable whereas CO2 emissions influence on mask use is working via a quadratic form; in other words, only highly polluted countries are characterized by a positive influence of pollution level (level of CO2 emissions in natural logarithm) on the percentage of population wearing a face mask.

These partial regressions give information about the potential socioeconomic determinants but need to be cautiously analysed due to the presence of potential issues, especially omitted variable bias and serial correlation issues.

##### 3.2.1 Partial linear regressions: observations on July, 15, 2020

Table 13: Masks determinants on July, 15, 2020: partial linear regressions (part A)

| | (1) | (2) | (3) | (4) | (5) | (6) |
|---|---|---|---|---|---|---|
| VARIABLES | masks | masks | masks | masks | masks | masks |
| co2 (log) | | -0.324*** | | | | |
| | | (0.107) | | | | |
| $co2^2 (log)$ | | 0.0157*** | | | | |
| | | (0.00508) | | | | |
| density (log) | 0.0432** | | | | | |
| | (0.0197) | | | | | |
| stringency | | | 0.00448*** | | | |
| | | | (0.00126) | | | |
| diabete | | | | 0.0657*** | | |
| | | | | (0.0247) | | |
| $diabete^2$ | | | | -0.00266** | | |
| | | | | (0.00103) | | |
| lgdp17 | | | | | -0.0363 | |
| | | | | | (0.0283) | |
| age652017 | | | | | | -0.00951** |
| | | | | | | (0.00426) |
| Constant | 0.395*** | 2.207*** | 0.298*** | 0.259** | 0.932*** | 0.681*** |
| | (0.0946) | (0.546) | (0.0891) | (0.126) | (0.268) | (0.0410) |
| Observations | 89 | 89 | 87 | 90 | 89 | 90 |
| R-squared | 0.056 | 0.073 | 0.150 | 0.065 | 0.021 | 0.072 |

Robust standard errors in parentheses
*** p<0.01, ** p<0.05, * p<0.1

Table 14: Masks determinants on July, 15, 2020: partial linear regressions (part B)

| | (1) | (2) | (3) | (4) | (5) | (6) |
|---|---|---|---|---|---|---|
| VARIABLES | masksfinal | masksfinal | masksfinal | masksfinal | masksfinal | masksfinal |
| gov_effectiveness_2018 | -0.0572 | | | | | |
| | (0.0354) | | | | | |
| pct_overweight | | -0.00328** | | | | |
| | | (0.00142) | | | | |
| altruism | | | 0.0523 | | | |
| | | | (0.0818) | | | |
| tolerance | | | | 0.0524*** | | |
| | | | | (0.0160) | | |
| $tolerance^2$ | | | | -0.000467*** | | |
| | | | | (0.000121) | | |
| cum cases0707 | | | | | 8.301** | |
| | | | | | (3.502) | |
| cum cases0906 | | | | | | 7.499 |
| | | | | | | (4.513) |
| Constant | 0.591*** | 0.755*** | 0.594*** | -0.796 | 0.556*** | 0.566*** |
| | (0.0239) | (0.0705) | (0.0290) | (0.527) | (0.0294) | (0.0280) |
| Observations | 87 | 86 | 59 | 52 | 88 | 88 |
| R-squared | 0.054 | 0.047 | 0.007 | 0.380 | 0.027 | 0.010 |

Robust standard errors in parentheses
*** p<0.01, ** p<0.05, * p<0.1

###### 3.2.2 Partial linear regressions: observations on April, 23-25, 2020

Table 15: Masks determinants on April, 23-25, 2020: partial linear regressions (part B)

| | (1) | (2) | (3) | (4) | (5) | (6) |
|---|---|---|---|---|---|---|
| VARIABLES | masks | masks | masks | masks | masks | masks |
| co2(log) | | -6.345 | | | | |
| | | (5.474) | | | | |
| $co2^2(log)$ | | 0.419 | | | | |
| | | (0.269) | | | | |
| density (log) | 0.754 | | | | | |
| | (0.969) | | | | | |
| stringency | | | 0.150** | | | |
| | | | (0.0728) | | | |
| diabete | | | | 1.752 | | |
| | | | | (1.249) | | |
| $diabete^2$ | | | | -0.0870* | | |
| | | | | (0.0476) | | |
| gdp (log) | | | | | -0.578 | |
| | | | | | (1.150) | |
| age65 | | | | | | 0.0929 |
| | | | | | | (0.215) |
| Constant | 14.83*** | 36.08 | 8.960** | 10.73 | 23.52** | 17.03*** |
| | (4.701) | (27.05) | (4.315) | (6.458) | (10.97) | (2.387) |
| Observations | 89 | 89 | 87 | 90 | 89 | 90 |
| R-squared | 0.005 | 0.120 | 0.052 | 0.019 | 0.002 | 0.002 |

Robust standard errors in parentheses
*** p<0.01, ** p<0.05, * p<0.1

Table 16: Masks determinants on April, 23-25, 2020: partial linear regressions (part B)

| VARIABLES | (1) masks | (2) masks | (3) masks | (4) masks |
|---|---|---|---|---|
| gov_effectiveness_2018 | -1.317 | | | |
| | (1.315) | | | |
| pct_overweight | | -0.0388 | | |
| | | (0.0783) | | |
| altruism | | | -7.081 | |
| | | | (5.463) | |
| tolerance | | | | 2.061* |
| | | | | (1.219) |
| $tolerance^2$ | | | | -0.0187** |
| | | | | (0.00904) |
| Constant | 18.66*** | 20.48*** | 20.29*** | -29.76 |
| | (1.482) | (3.944) | (1.943) | (39.19) |
| Observations | 87 | 86 | 59 | 52 |
| R-squared | 0.009 | 0.002 | 0.027 | 0.188 |

Robust standard errors in parentheses
*** p<0.01, ** p<0.05, * p<0.1

##### 3.3 Effect of education: observations on July, 15, 2020

We have tested the effect of education using the last available data about the schooling variable from the World Bank and also the last PISA score. We expect that a better education level enhance the mask wearing as well as hygiene and compliance in a general way.

##### 3.4 Effect of overweight population: observations on July, 15, 2020

We have tested the effect of education using the last available data about the overweight proportion variable.

##### 3.5 Additional multivarable regressions: controlling for non-pharmaceutical measures

We control for the effect of other mitigation measures on mask wearing on July, 15, 2020 by introducing four dummy variables in our regression model: travel restriction (*travel*), testing policy (*testing*), school closures policy (*school*), surveillance and tracking policy (*surveillance*). All data come from the Porcher Simon database at https://response2covid19.org/.

##### 3.6 Additional multivarable regressions: controlling for trust in government

Some studies assume that differences in government and politicians trust can impact the effectiveness of control policies. We computed the proportion of people that have 'not trust at all' in their government from World Value Survey databese (2017-2020 survey). A high proportion of people that do not trust in government is not associated to a reduction of mask wearing proportion. However, note that the number of observation is limited and results need to be cautiously interpreted. We can not completely confirm that people that do not trust at all are the mask offenders and explain heterogeneities about mask wearing across countries.

Table 17: Education effect estimates

| VARIABLES | (1) masksfinal | (2) masksfinal |
|---|---|---|
| ldensity2017 | 0.0367** | 0.0917*** |
| | (0.0182) | (0.0216) |
| lco2014 | -0.150 | -0.953* |
| | (0.156) | (0.506) |
| lco2014sq | 0.00778 | 0.0427* |
| | (0.00691) | (0.0215) |
| cumulatedcases0707pop100 | 0.0182 | 0.0741 |
| | (0.0238) | (0.107) |
| stringency | 0.00601*** | 0.00647* |
| | (0.00124) | (0.00345) |
| school2017 | -0.00342 | |
| | (0.00338) | |
| pisa | | -0.00117 |
| | | (0.00122) |
| Constant | 1.079 | 5.542* |
| | (0.721) | (3.095) |
| Observations | 74 | 33 |
| R-squared | 0.368 | 0.524 |

Robust standard errors in parentheses
*** p<0.01, ** p<0.05, * p<0.1

Table 18: Overweight effect estimates

| VARIABLES | (1)<br>masksfinal | (2)<br>masksfinal |
|---|---|---|
| density | | 0.0278 |
| | | (0.0212) |
| co2 | | -0.304*** |
| | | (0.101) |
| $co2^2$ | | 0.0148*** |
| | | (0.00477) |
| cum cases0707 | | 0.102** |
| | | (0.0400) |
| diabete | | 0.00978 |
| | | (0.00638) |
| pct_overweight | 0.0199* | 0.0285*** |
| | (0.0108) | (0.00938) |
| $pct\_overweight^2$ | -0.000261** | -0.000352*** |
| | (0.000124) | (0.000105) |
| Constant | 0.323 | 1.437** |
| | (0.206) | (0.546) |
| Observations | 86 | 83 |
| R-squared | 0.084 | 0.244 |

Robust standard errors in parentheses
** 4* p<0.01, ** p<0.05, * p<0.1

Table 19: Estimates with mitigation policies controls

| | (1) | (2) | (3) |
|---|---|---|---|
| VARIABLES | masks | masks | masks |
| density | | 0.0436** | 0.0455** |
| | | (0.0182) | (0.0186) |
| co2 | | -0.365*** | -0.365*** |
| | | (0.111) | (0.113) |
| $co2^2$ | | 0.0172*** | 0.0173*** |
| | | (0.00539) | (0.00550) |
| stringency | | 0.00485*** | 0.00500*** |
| | | (0.00120) | (0.00122) |
| cum cases0707 | | | 0.0286 |
| | | | (0.0267) |
| testing | -0.0116 | 0.0575 | 0.0814 |
| | (0.0673) | (0.0555) | (0.0551) |
| surveillance | -0.0239 | 0.0234 | 0.0153 |
| | (0.0907) | (0.0630) | (0.0625) |
| school | -0.242 | -0.188 | -0.176 |
| | (0.210) | (0.122) | (0.123) |
| travel | 0.102 | 0.0228 | 0.0343 |
| | (0.0951) | (0.0801) | (0.0826) |
| Constant | 0.507*** | 1.947*** | 1.900*** |
| | (0.0910) | (0.553) | (0.558) |
| Observations | 89 | 84 | 82 |
| R-squared | 0.037 | 0.333 | 0.363 |

Robust standard errors in parentheses
*** p<0.01, ** p<0.05, * p<0.1

Table 20: Estimates with trust in government variable

| | (1) | (2) |
|---|---|---|
| VARIABLES | masksfinal | masksfinal |
| density | 0.0512* | 0.0640* |
| | (0.0262) | (0.0350) |
| co2014 | -0.0852 | -0.119 |
| | (0.406) | (0.419) |
| co2sq | 0.00698 | 0.00835 |
| | (0.0173) | (0.0178) |
| stringency | 0.00534*** | 0.00516*** |
| | (0.00127) | (0.00130) |
| cum cases0707 | | 0.0401 |
| | | (0.101) |
| government_confidence | 0.00265 | 0.00293 |
| | (0.00170) | (0.00182) |
| Constant | -0.000191 | 0.143 |
| | (2.384) | (2.446) |
| Observations | 45 | 44 |
| R-squared | 0.416 | 0.422 |

Robust standard errors in parentheses
*** p<0.01, ** p<0.05, * p<0.1

### 4 Definitions and data sources

#### 4.1 Panel data set

For each variable, we indicate the acronym, the definition and the sources respectively.

- *casepop* and *deathpop*

Data have been collected from Johns Hopkins University on July 15, 2020. Covid-19 cases and deaths on a daily basis reported by John Hopkins University. We compute infected cases and fatality rates by using the cumulative values divided by the total population of country $i$ to obtain a *percapita* ratio at a daily frequency. Our panel data are available from January, 1, 2020 to July, 15, 2020. The *casepop* variable is computed as the following ratio: number of infected cases on day d over total population of the country. The *deathpop* variable is computed as the fatality rate: number of cumulative fatalities on day $d$ over total population of the country $i$. To take into account the dynamics of the epidemic, the incubation period (time between $t$ and $t-k$) and other delays, we also use lagged $casepop(t-k)$ and $deathpop(t-k)$ variables.

- $new_tests$

This variable denotes the number of new Covid-19 tests at a daily frequency per thousand variable comes from Hasell, J., Mathieu, E., Beltekian, D. et al. A cross-country database of COVID-19 testing. Sci Data 7, 345 (2020).

- *masks*

The data come from the survey from Maryland University jointly conducted with Facebook: Barkay, N. et al., Weights and Methodology Brief for the COVID-19 Symptom Survey by University of Maryland and Carnegie Mellon University, in Partnership with Facebook", arXiv e-prints, (2020). It consists in aggregate weighted estimates masks wearing variable is computed as the number of individuals reporting a mask use on the Facebook plateform. As explained by the author: "we are inviting Facebook app users in more than 200 countries or territories globally to take a survey collected by faculty at the University of Maryland (UMD) Joint Program in Survey Methodology (JPSM). As part of this initiative, we are applying best practices from survey statistics to design and execute two components: (1) sampling design and (2) survey weights, which make the sample more representative of the general population". "We provide weights for two sets of sample respondents separately for both the CMU US and UMD global surveys. First, we provide weights for respondents who answered the questions needed to calculate the aggregate estimates of COVID-like Illness (CLI) reported in the CMU and UMD APIs. Second, we provide weights for a larger set of respondents who answered a minimum of two questions in the surveys." We extracted data from Maryland University link at https://covidmap.umd.edu/api.html. More precisely, we used the following $percent_mc$ variable: weighted percentage of survey respondents that have reported use mask cover. Using the survey data, the authors of the survey estimate the percentage of people in a given country or region, on a given day that use mask cover. The $smoothed_mc$ consisting in seven-day rolling average of $percent_m c$ values as well as $smoothed_m c_s e$ that is the standard error of $smoothed_m c$. has also been considered. We finally computed a standardized index: $smoothed_m c/smoothed_m c_s e$

- *stringency*

The Stringency Index summarizes the severity of the lockdown measures. The policy response tracker data has been developed by Oxford's public policy school, the Blavatnik School of Government. They publish it as the Coronavirus Government Response Tracker (OxCGRT). The OxCGRT researchers calculate a summary measure of nine of the response metrics called the Government Stringency Index: school closures; workplace closures; cancellation of public events; restrictions on public gatherings; closures of public transport; stay-at-home requirements; public information campaigns; restrictions on internal movements; and international travel controls. The specific policy and response categories are coded as follows and then each indicator is rescaled to get a general score between 0 and 100 (100 representing the highest degree of strictness).

School closures: 0 - No measures 1 - recommend closing 2 - Require closing (only some levels or categories, eg just high school, or just public schools) 3 - Require closing all levels No data - blank

Workplace closures: 0 - No measures 1 - recommend closing (or work from home) 2 - require closing (or work from home) for some sectors or categories of workers 3 - require closing (or work from home) all but essential workplaces (eg grocery stores, doctors) No data - blank

Cancel public events: 0- No measures 1 - Recommend cancelling 2 - Require cancelling No data - blank

Restrictions on gatherings: 0 - No restrictions 1 - Restrictions on very large gatherings (the limit is above 1000 people) 2 - Restrictions on gatherings between 100-1000 people 3 - Restrictions on gatherings between 10-100 people 4 - Restrictions on gatherings of less than 10 people No data - blank

Close public transport: 0 - No measures 1 - Recommend closing (or significantly reduce volume/route/means of transport available) 2 - Require closing (or prohibit most citizens from using it)

Public information campaigns: 0 -No COVID-19 public information campaign 1 - public officials urging caution about COVID-19 2 - coordinated public information campaign (e.g. across traditional and social media) No data - blank

Stay at home: 0 - No measures 1 - recommend not leaving house 2 - require not leaving house with exceptions for daily exercise, grocery shopping, and 'essential' trips 3 - Require not leaving house with minimal exceptions (e.g. allowed to leave only once every few days, or only one person can leave at a time, etc.) No data - blank

Restrictions on internal movement: 0 - No measures 1 - Recommend movement restriction 2 - Restrict movement

International travel controls: 0 - No measures 1 - Screening 2 - Quarantine arrivals from high-risk regions 3 - Ban on high-risk regions 4 - Total border closure No data - blank

Testing policy 0 – No testing policy 1 – Only those who both (a) have symptoms AND (b) meet specific criteria (eg key workers, admitted to hospital, came into contact with a known case, returned from overseas) 2 – testing of anyone showing COVID-19 symptoms 3 – open public testing (eg "drive through" testing available to asymptomatic people) No data

Contract tracing 0 - No contact tracing 1 - Limited contact tracing - not done for all cases 2 - Comprehensive contact tracing - done for all cases No data

Face coverings 0- No policy 1- Recommended 2- Required in some specified shared/public spaces outside the home with other people present, or some situations when social distancing not possible 3- Required in all shared/public spaces outside the home with other people present or all situations when social distancing not possible 4- Required outside the home at all times regardless of location or presence of other people

- *temperatures* conditions consist in mean daily temperatures (in Celcius degrees) from daily have been aggregated by Aviskar Bhoopchand, Andrei Paleyes, Kevin Donkers, Nenad Tomasev, Ul- rich Paquet (2020), DELVE Global COVID-19 Dataset and are available at http://rs-delve.github.io/data /global-dataset.html. For each country, all climatic observations have been weighted by the population. In addition, the temperature variable has been standardized (divided by standard deviation) for comparisons purposes and for coefficients scale homogeneity.

- *mobility*

Finally, we use Apple mobility trend reports data that are available at: https://covid19.apple.com/mobility. The database consists in two indicators: $Mobility_travel_driving$ and *walking*. Both indicators are indexes (January, 13, 2020 as the 100 basis) and are computed as the % change since the January, 13, 2020 basis. Apple's mobility trend reports show how human mobility has changed in countries and cities worldwide since January 2020 and are based on location data of Apple's "maps" services. It is designed to help mitigate the spread of COVID-19, provide governments, research institutions, health authorities, and the general public with insights on the effects on human mobility of national and regional lockdown policies. The data covers 63 countries. All data are shared on an aggregated level and Apple does not keep a history of users' mobility behaviour. Data from Google reports have also been considered as an alternative. We proceed to some tests and the dynamics from both databases are very similar. Since Apple data are available on a more longer period (January, 13, 2020 versus February, 15, 2020), we have chosen Apple data in our paper.

#### 4.2 Cross-sectional data set

For each variable, we indicate the acronym, the definition and the sources respectively.

- *age*65

Population aged over 65 as a percentage of the total population from the World Bank Database. Population is based on the de facto definition of population, which counts all residents regardless of legal status or citizenship.

- *density*

Population density from the World Bank Database. is midyear population divided by land area in square kilometers. Population is based on the de facto definition of population, which counts all residents regardless of legal status or citizenship–except for refugees not permanently settled in the country of asylum, who are generally considered part of the population of their country of origin. Land area is a country's total area, excluding area under inland water bodies, national claims to continental shelf, and exclusive economic zones. In most cases the definition of inland water bodies includes major rivers and lakes.

- $CO2$ dioxide (CO2) emissions from the World Bank Database are those stemming from the burning of fossil fuels and the manufacture of cement. They include carbon dioxide produced during consumption of solid, liquid, and gas fuels and gas flaring. The natural logarithm of the CO2 emissions has been considered in our econometric regressions.

- *Diabetic*

Diabete measures Diabetes prevalence refers to the percentage of people ages 20-79 who have type 1 or type 2 diabetes complied by the World Bank Database (International Diabetes Federation, Diabetes Atlas).

- *Overweight*

Overweight measures the percentage of the adult population which is overweighted.

*GDP*

GDP measures GDP per capita in purchasing power parity for 2017 from the World Bank Database.

*School*

School enrollment from the World Bank Database., primary, $\%istheratiooftotalenrollment, regardlessofag$

*PISA*

The Reading performance (PISA) Boys / Girls, Mean score in 2018, has been considered in our study. Source: PISA: Programme for International Student AssessmentData consist in https://www.oecd.org/pisa/data/

- *altruism*

Altruism from Falk et al. (2018) was measured through a combination of one qualitative and one quantitative item, both of which are related to donations. The qualitative question asked respondents how willing they would be to give to good causes without expecting anything in return on an 11-point scale. The quantitative scenario depicted a situation in which the respondent unexpectedly received 1,000 euros and asked them to state how much of this amount they would donate (Table I).

- *tolerance*

We used the tolerance intentions index from the World Value Survey database. More precisely, we selected answers to the question 12 untitled 'Tolerance and respect for other people' from the survey 2017-2020. Question wording 'Here is a list of qualities that children can be encouraged to learn at home. Which, if any, do you consider to be especially important? Please choose up to five'. Our index has been computed by reporting the percentage (over the total of responses) of 'important' responses.

- $government\_confidence$

We also tested the possibility that trust in politicians and government can impact the compliance in line with (5). We used data about the question 71 untitled 'Confidence: The government' from the survey 2017-2020 (World Value Survey database). We computed the percentage score concerning the "not at all" answer. In our opinion, it is a mean to capture the proportion of people in a given country that do not trust at all in the government and public guidance.

#### 5 List of countries

Considering available data, 96 countries have been finally considered in our statistical analysis:

Afghanistan Albania Algeria Angola Argentina Armenia Australia Austria Azerbaijan Bangladesh Belarus Belgium Bolivia Bosnia and Herzegovina Brazil Bulgaria Burkina Faso Cambodia Cameroon Canada Chile Colombia Costa Rica Croatia Cyprus Czechia Denmark Dominican Republic Ecuador Egypt Salvador Estonia Ethiopia Finland France Georgia Germany Ghana Greece Guatemala Honduras Hong Kong Hungary Iceland India Indonesia Iran Iraq Ireland Israel Italy Jamaica Japan Jordan Kazakhstan Kenya Kuwait Kyrgyzstan Laos Libya Lithuania Madagascar Malaysia Mali Mexico Moldova Morocco Nepal Netherlands New Zealand Nicaragua Nigeria Norway Oman Pakistan Panama Paraguay Peru Philippines Poland Portugal Qatar Republic of Korea Romania Russia Saudi Arabia Serbia Singapore Slovakia Slovenia South Africa Spain Sri Lanka Sudan Sweden Switzerland Taiwan Tanzania Thailand Tunisia Turkey Ukraine United Arab Emirates United Kingdom United States of America Uruguay Uzbekistan Venezuela Vietnam
